# Whether the Weather Will Help Us Weather the COVID-19 Pandemic: Using Machine Learning to Measure Twitter Users’ Perceptions

**DOI:** 10.1101/2020.07.29.20164814

**Authors:** Marichi Gupta, Aditya Bansal, Bhav Jain, Jillian Rochelle, Atharv Oak, Mohammad S. Jalali

## Abstract

**Objective:** The potential ability for weather to affect SARS-CoV-2 transmission has been an area of controversial discussion during the COVID-19 pandemic. Individuals’ perceptions of the impact of weather can inform their adherence to public health guidelines; however, there is no measure of their perceptions. We quantified Twitter users’ perceptions of the effect of weather and analyzed how they evolved with respect to real-world events and time.

**Materials and Methods:** We collected 166,005 tweets posted between January 23 and June 22, 2020 and employed machine learning/natural language processing techniques to filter for relevant tweets, classify them by the type of effect they claimed, and identify topics of discussion.

**Results:** We identified 28,555 relevant tweets and estimate that 40.4% indicate uncertainty about weather’s impact, 33.5% indicate no effect, and 26.1% indicate some effect. We tracked changes in these proportions over time. Topic modeling revealed major latent areas of discussion.

**Discussion:** There is no consensus among the public for weather’s potential impact. Earlier months were characterized by tweets that were uncertain of weather’s effect or claimed no effect; later, the portion of tweets claiming some effect of weather increased. Tweets claiming no effect of weather comprised the largest class by June. Major topics of discussion included comparisons to influenza’s seasonality, President Trump’s comments on weather’s effect, and social distancing.

**Conclusion:** There is a major gap between scientific evidence and public opinion of weather’s impacts on COVID-19. We provide evidence of public’s misconceptions and topics of discussion, which can inform public health communications.

## INTRODUCTION

### Background and Significance

Since the beginning of the outbreak, one of the major questions has been whether the transmission of SARS-CoV-2 is seasonal, such as with influenza,^1^ MERS,^2^ or SARS.^3^ While there was limited research and consensus at the beginning of the pandemic on the impact of weather and seasonality on the transmission of SARS-CoV-2,^4-12^ a growing body of evidence has suggested that the effect of weather conditions is modest and that weather alone is not sufficient to quench the pandemic.^13^ Despite (limited) academic consensus, what the public thinks is unknown, which motivated our research.

As COVID-19 has disrupted the global population, many have turned to social media platforms such as Twitter to navigate COVID-19. While Twitter’s effectiveness at disseminating information can be leveraged to share public health information for social good, it can also promote misinformation.^14^ As the virus continues to spread, chatter online has increased in volume, and one particularly contentious topic of discussion surrounds the myth that heat can effectively kill the virus.^15^ While it is not uncommon for public opinion to contradict scientific literature, the continuous debate, uncertainty, and lack of consensus among experts exacerbated this specific public misconception.^16,17^ As public comments are good predictors for individual’s behaviors, measuring and analyzing the social perception of the weather’s impact on COVID-19 may help predict adherence to public health policy and guidelines.

### Objectives

This study examined Twitter users’ perceptions concerning the weather’s effect on the spread of COVID- 19 with natural language processing and machine learning techniques. Specifically, the research objectives were to identify: (1) the perceived impact of weather in relevant tweets and classify them accordingly, and (2) if and how these perceptions changed throughout the pandemic. To investigate these, we trained a support vector machine classifier to measure what proportion of tweets claim there is an effect of weather, and exhibit time-series trends for a subset of relevant tweets. To detect perceptions outside of this effect-oriented framework, we employed unsupervised learning to discover unexpected discussion topics.

This study is one of many to use machine learning and natural language processing to retrieve information about public perception through social media for public health purposes,^18,19^ but the first to study the perception of the weather’s impact on COVID-19. We hope that this work can inform public policy and research as the COVID-19 pandemic response continues.

## MATERIALS AND METHODS

### Tweet Collection

Using Twitter’s Premium application programming interface (API) for historical search, we collected 166,005 tweets from January 23 to June 22, 2020 with the query “(coronavirus OR covid OR covid19) AND weather.” This query checked all tweet components for a match, including the tweet’s text, the text of any attached articles or media, and any URL text included with the tweet. We only collected quoted or original tweets, not retweets, that were written in English. We also did not limit the data to any specific location. For tweets replying to or quoting another tweet, we fetched the text of the other tweet. For tweets sharing an article, we collected the article headline and description as displayed on Twitter. The tweet text, article data, and any replied-to/quoted tweets were then merged for analysis. Figure 1 presents our research method and the flow of its processes, which are discussed below.

**Figure 1:**
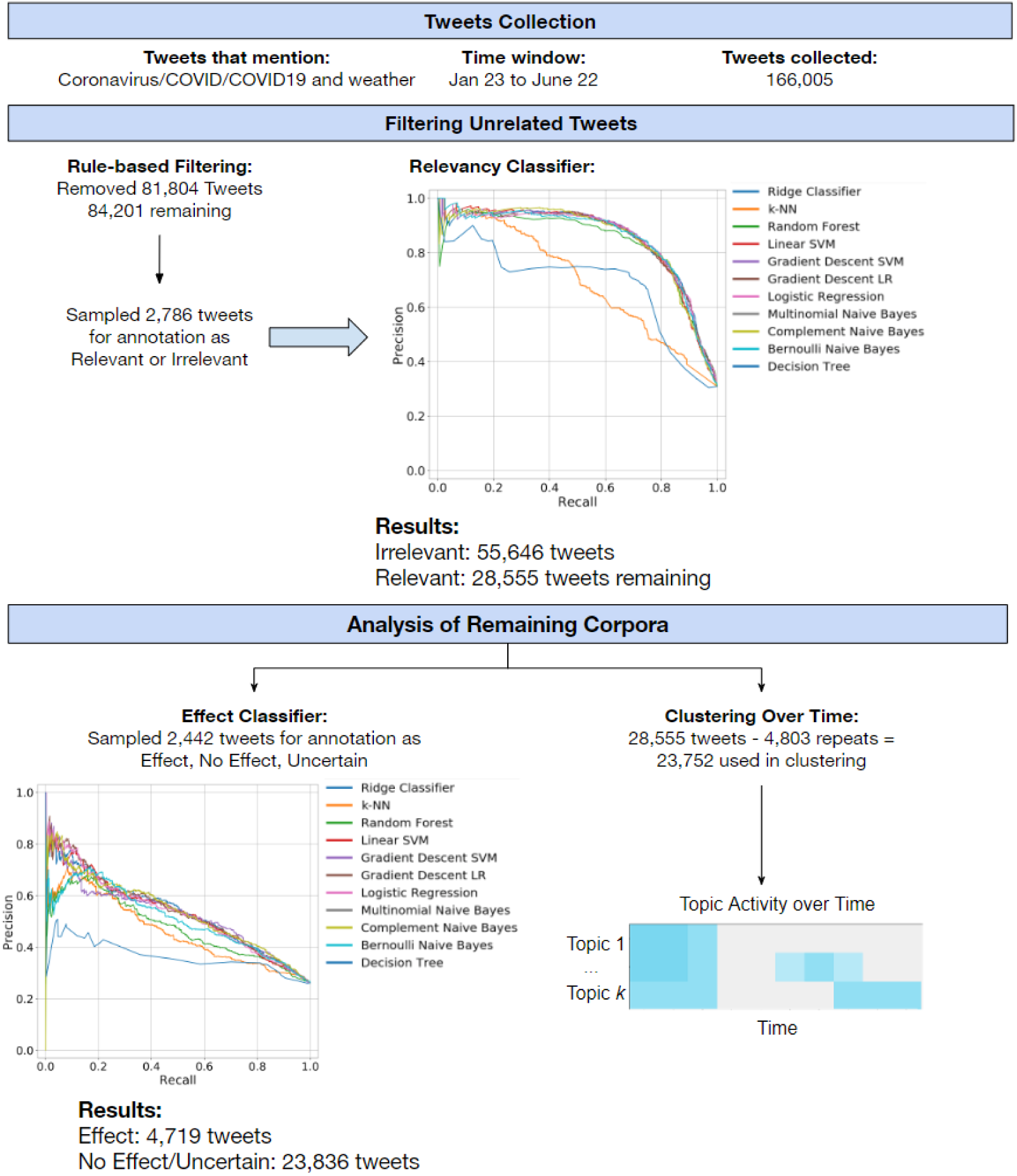
Flow diagram of filtering and machine learning processes.

### Reducing Corpus to Relevant Tweets

#### Rule-based Filtering

Initially, we cleaned tweets by removing any non-alphanumeric characters (including emojis), mentions of other users, and hashtags at the end of the tweet, and then we further standardized with lemmatization and stemming (see Supplementary S2 for more details). Following common techniques used for social media analysis in other domains,^20^ we employed rule-based filtering to narrow our corpus down and remove noise. The rule-based filtering consisted of three rules applied sequentially. First, we filtered out false positives coming from the sheer popularity of our keywords (e.g., a tweet commenting on pleasant weather and ending with “#coronavirus”) and removed tweets where the keywords were split across different parts of the tweet (e.g., “weather” only appearing in the article text, and “covid” only in the tweet itself). Second, we discarded tweets using “weather” as a verb or idiomatically (e.g., “under the weather”). Finally, we restricted the tweets to those posted by individuals, not news organizations, since individual perception was the focus of study. The strengths of these three rules were verified manually (see S3).

#### Relevancy Classification

We used machine learning to further reduce the corpus to tweets that had insightful relationships between weather and the spread of COVID-19.

##### Annotation

To create training data for the classifier, two annotators (JR, BJ) labeled a set of tweets based on pre- defined inclusion criteria, which defined a tweet as relevant if it referenced a causal or correlative relation between weather and coronavirus spread, and irrelevant otherwise. Tweets presenting a causal relationship declared the weather to have a direct impact on the spread of COVID-19 (e.g., high temperatures killing the virus) while a correlative relationship declared an indirect impact (e.g., reduced social distancing during pleasant weather). Irrelevant tweets mentioned weather and COVID-19 but did not establish a relationship between them (e.g., extreme weather causing additional strain in hard-hit areas). Annotators marked a shared pilot set of 100 tweets to calibrate on these criteria. After resolving any discrepancies, annotators labeled a full set of training data for our machine learning classifiers.

##### Natural Language Processing and Featurization

Text featurization was used to convert tweets into meaningful vectors for machine learning analysis. Three vectorization techniques were used: Bag of Words (BOW), Term Frequency-Inverse Document Frequency (TF-IDF), and Embeddings from Language Models (ELMo), a state-of-the-art technique that utilizes word embeddings.^21^ ELMo factors in the surrounding context for each word (i.e., the words around it) for its vectorization, while BOW and TF-IDF do not.^22^ For BOW and TF-IDF, we removed stop words (set of commonly used words which do not contribute to the context of the tweet) and also words that only appeared in 1% of all tweets or less.

We tested 11 classification models for performance on relevancy classification: Ridge Classifier, Logistic Regression, k-Nearest Neighbors, Support Vector Machine, Logistic Regression with Gradient Descent, Support Vector Machine with Gradient Descent, Multinomial Naïve Bayes, Complement Naïve Bayes, Bernoulli Naïve Bayes, Random Forest Classifier, and Decision Trees (see S5). We used Scikit-learn’s machine learning libraries.^23^

We performed a five-fold outer cross-validation on our training dataset to select the optimal model with five-fold inner cross-validation to find the ideal hyperparameters (see S4). For each of our models, we evaluated and reported the Area under the Precision-Recall curve (AUC-PR) and Area under the Receiver Operating Characteristic curve (AUC-ROC)—for definitions, see^24^. Both metrics are presented, but we chose to optimize with respect to AUC-PR since it provides a better assessment of model performance for imbalanced datasets, where AUC-ROC be overly optimistic.^24-26^ We took the best performing model to be our “Relevancy Classifier” that produced the corpus for analysis, both for the claimed effect of weather and for topics of discussion.

### Analyzing Tweets for Effect

To classify tweets based on the type of effect the user expected the weather to have on the spread of COVID-19, we trained another machine learning classifier.

#### Effect Classification

##### Annotation

We first annotated a new batch of tweets (distinct from the relevancy annotation set) based on if they claimed weather to have some effect and used this as training data. After calibrating on a pilot set of 200 tweets, annotators (JR, BJ, MG) first labeled tweets into one of three categories: “effect,” where the tweet suggested that weather had an impact on COVID-19; “no effect,” where the tweet suggested weather had no impact; and “uncertain,” where the tweet was uncertain to the effect or made no clear claim to an effect.

Additionally, within the “effect” category, tweets were labeled based on whether the tweet suggested COVID-19 would: i) improve with warmer weather, ii) worsen with warmer weather, iii) improve with cooler weather, or iv) worsen with cooler weather. This class scheme assumed that temperature was the key driver of discussions; we found this to be representative of discussion on Twitter as well as the main focus of academic literature on the weather’s impact.^4,5,7,8^ The inclusion, for instance, of both “improve with warmer weather” and “worsen with cooler weather” was to avoid any assumption of a linear effect of temperature given that non-linear effects have been documented.^13^

For qualitative analysis, the annotators recorded the mechanisms users reported for the weather’s impact on coronavirus spread, such as sunlight destroying the virus. These mechanisms provided insight into the theories of the weather’s impact being discussed and are reported in the Discussion.

##### Natural Language Processing and Featurization

For our Effect Classifier, the same machine learning techniques were used from our Relevancy Classifier (as described above) with one modification: for the trinary classification, we optimized with respect to balanced accuracy, since AUC-PR and AUC-ROC do not extend to multiclass problems.

### Analyzing Tweets for Topic via Clustering

To extract unexpected topics of discussion, we performed unsupervised learning to cluster the tweets and determined topics through inspection of the clusters. After removing repeated tweets (not retweets) and attached article data, we used *k*-means clustering to group tweets into *k* clusters—other methods, specifically *k-*medoids and latent Dirichlet allocation^27^ were also explored (see S7). Clustering was performed on the same TF-IDF vectors generated for effect analysis, and cluster sizes in *k*=10, 15, 20, 25, and 30 were tested. Each cluster was associated with an output of the top 20 keywords, based on highest TF-IDF scores. Outputs from each of the clustering configurations were inspected manually for the cohesiveness of topics.

## RESULTS

### Data Preparation and Annotation

The data pipeline is displayed in Figure 1, with inspiration taken from Ong et al.^28^ Overall, rule-based filtering reduced the corpus from 166,005 to 84,201 tweets. For relevancy classification, annotators labeled a random sample of 2,786 tweets, and the Relevancy Classifier was trained on this. Then, for effect classification, the “effect” of a random sample of 2,442 relevant tweets (out of 28,555) was annotated per the Effect Class and annotation scheme introduced earlier, with results shown in Table 1.

**Table 1:** Manual Annotation Scheme for Effect and Class Proportions

| Class | Proportion (out of 2,442) |
| --- | --- |
| Uncertain | 40.4% (987) |
| No Effect | 33.5% (817) |
| Effect | 26.1% (638) |
| Improve Warmer Weather | 585 |
| Worsen Warmer Weather | 33 |
| Improve Cooler Weather | 4 |
| Worsen Cooler Weather | 16 |

### Relevancy Classification Using Machine Learning

Our relevancy classifier identified tweets discussing the weather’s impacts on COVID-19, with the volumes over time shown in Figure 2. Three example peaks in activity are shown in the figure along with the most commonly shared headline in the dataset from that day (more details are available in S6). The best performing classifier for this phase of learning was Gradient Descent Support Vector Machine with TF-IDF featurization, with AUC-PR (95% CI)=0.862 (0.853, 0.871) and AUC-ROC (95% CI)=0.916 (0.907, 0.925).

**Figure 2:**
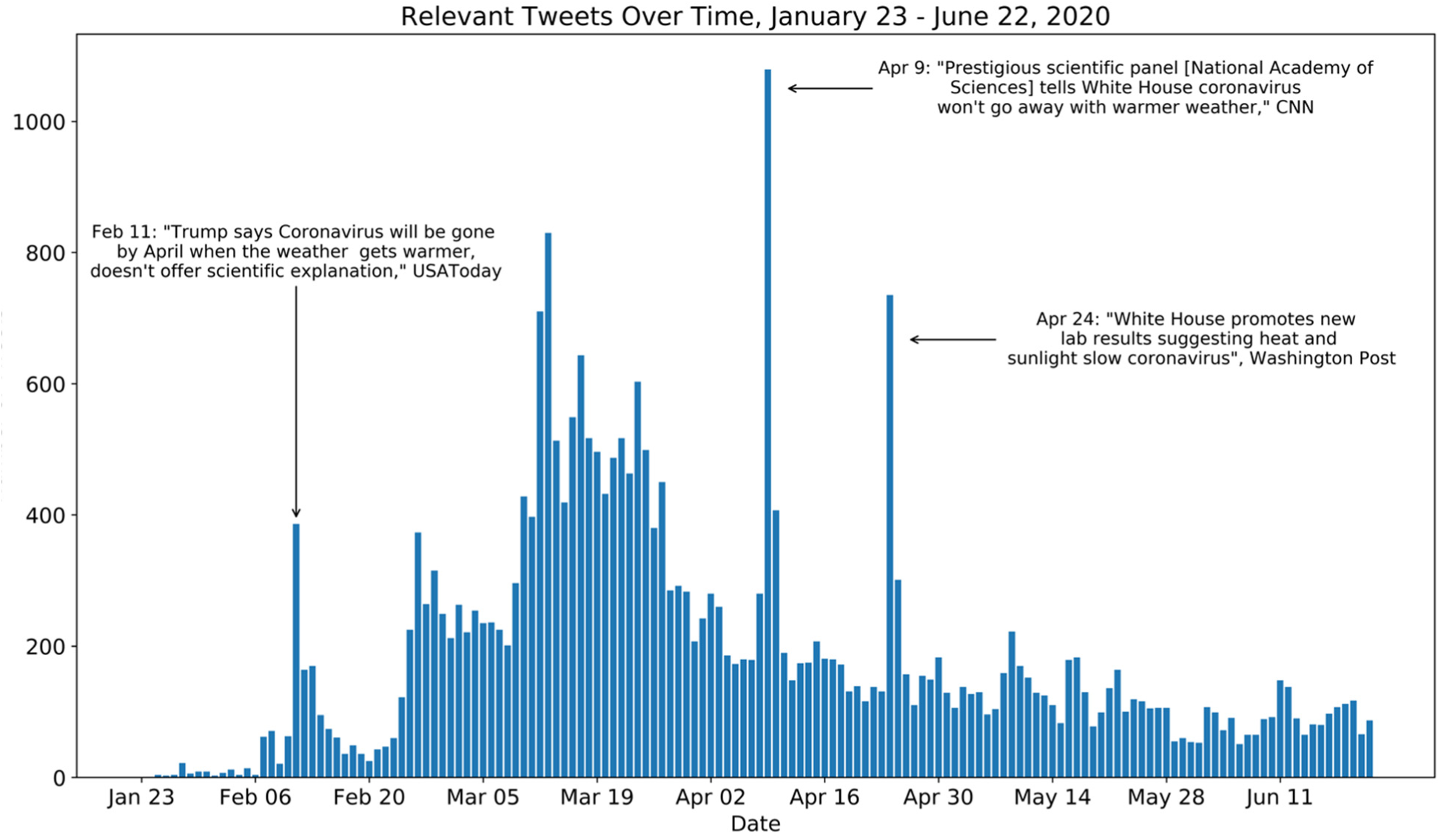
Relevant original tweet volumes over time, with most frequent headlines and reporting organizations on three key peaks identified.

### Effect Analysis

#### Manual Annotation Results

The 2,442 annotated tweets were separated according to their effect label (effect, no effect, uncertain) and plotted in Figure 3.

**Figure 3:**
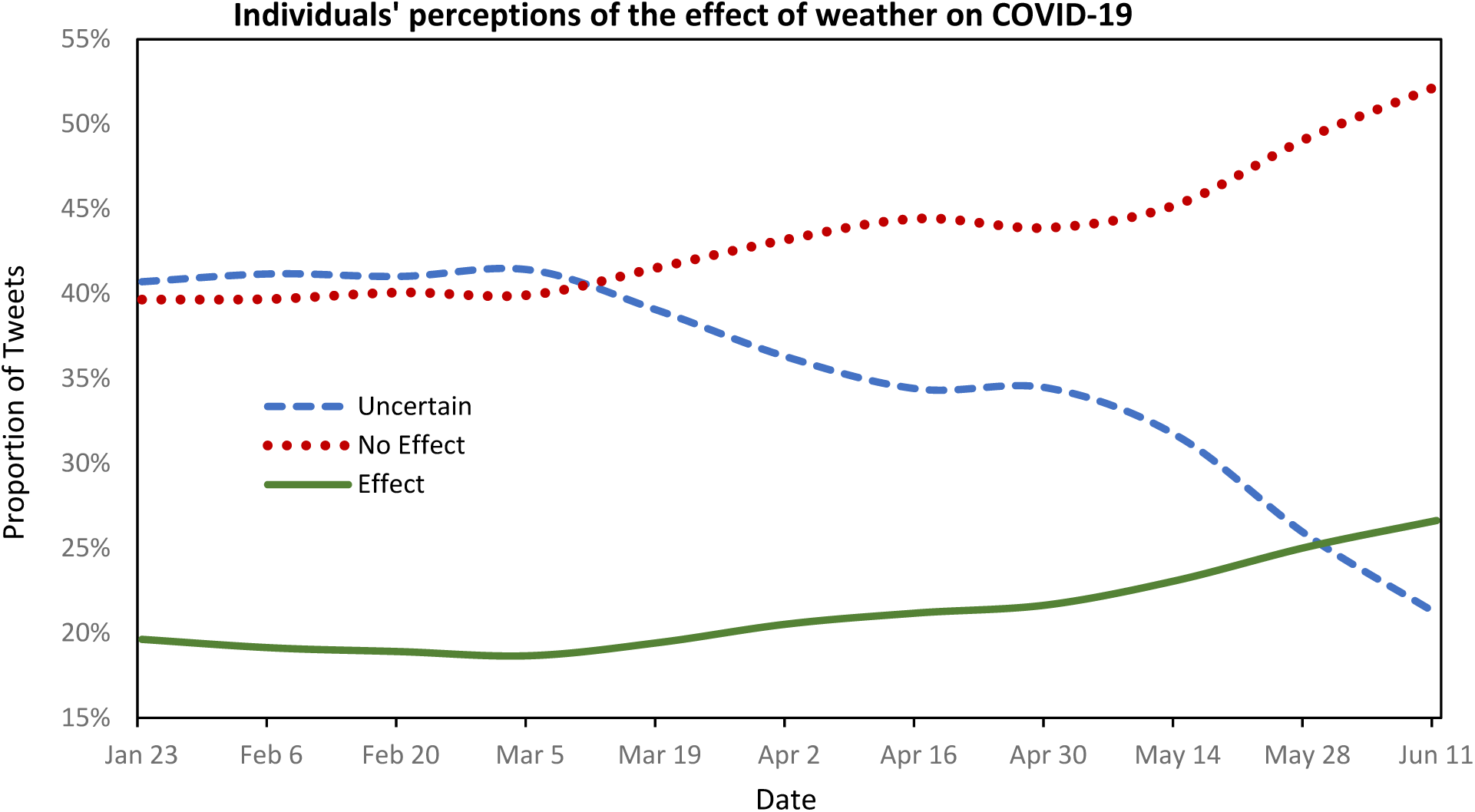
Class proportion over time for annotated Tweets. Tweets are smoothed by 7 days, binned in 14-day windows, and weighted according to the individual tweet’s number of retweets.

#### Results of Machine Learning Classifier

Using the manual annotations for our Effect Classifier, we attempted to predict the perception of a tweet according to the three classes. However, the multiclass scheme proved too difficult for machine to solve (see S5), but after collapsing our class scheme to a binary “effect” vs. “no effect/uncertain” (combining those two categories) the performance of the model improved (see Table 2 and S5). We still present these to show the machine did learn to identify effect to an extent, accomplishing our goal of identifying perception even after limiting our analysis to the coarser class scheme. The AUC-PR and AUC- ROC scores are reported in Table 2; for reference, a baseline classifier (one that randomly predicts the class) has an AUC-PR of 0.261—the proportion of the “effect” class in Table 1– and an AUC-ROC of 0.5.

**Table 2:**
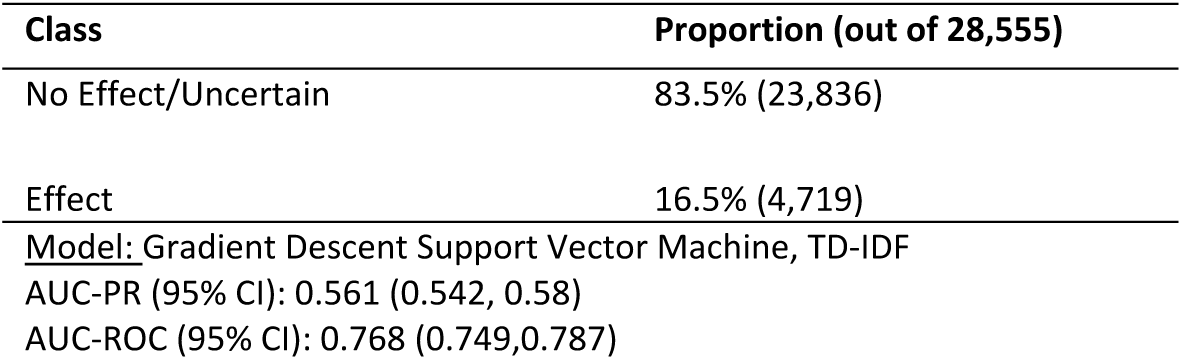
Machine learning classification results

### Clustering

The optimal configuration for *k-*means clustering was *k*=25 to retrieve clear topics of discussion (see S7). After dropping 4,803 repeated tweets, we clustered on 23,752 tweets. Twenty-four of the assigned clusters produced clearly delineated topics, while the remaining cluster was vague and contained general comments about weather and coronavirus.

Figure 4 displays a heatmap tracking discussion frequency across ten selected topics over time. Boxes in the heatmap are shaded only for weeks where a topic exceeded its average level of discussion in the corpus, which allows for meaningful interpretation of when a topic is more active than usual.

**Figure 4:**
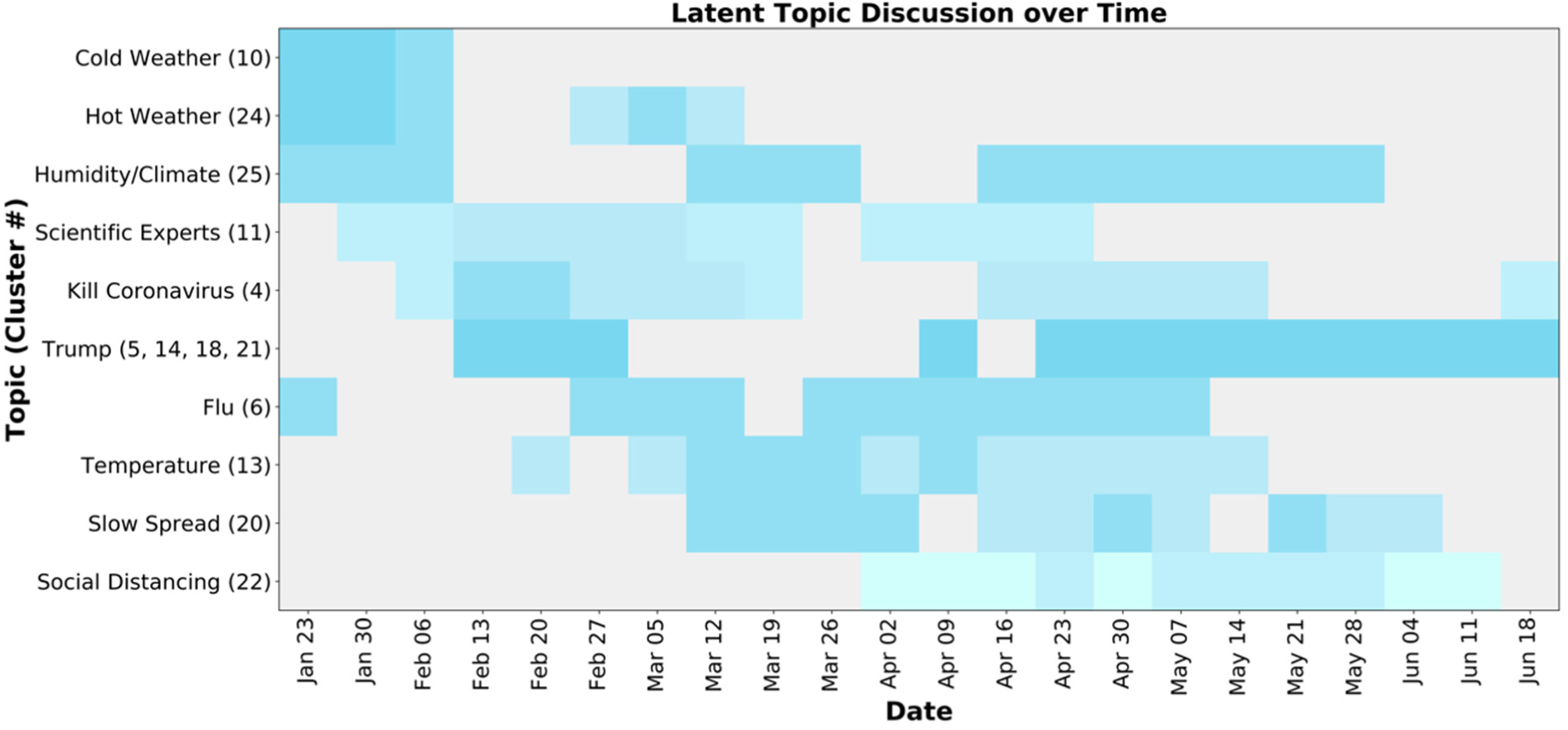
Cluster Frequencies over Time by Week, color coding presents the frequency of discussion, where darker blue is the highest frequency.

The ten clusters plotted in Figure 4 are particularly meaningful. Specifically, cluster 10 discussed the effect of cold weather on coronavirus spread; cluster 24 discussed the effect of hot weather on coronavirus spread; cluster 25 consisted of tweets proclaiming the relationship between different climates and general viral spread; cluster 11 discussed opinions propelled by scientific experts; cluster 4 focused on the ability of weather to ‘kill’ the coronavirus; clusters 5, 14, 18, and 21 referenced the Trump administration; cluster 6 included tweets comparing the coronavirus to influenza viruses; cluster 13 highlighted relationships between temperature and coronavirus spread; cluster 20 contained tweets considering the ability of weather to ‘slow spread’ of the virus; and cluster 22 consisted of conversation revolving around social distancing. (See S7 for the top 20 keywords, sample tweets, and proportions for each cluster.)

## DISCUSSION

Our analysis shows that Twitter user’s perception of the weather’s impact on the spread of COVID-19 varied greatly. Our results help quantify individuals’ perceptions and reveal central topics of discussion surrounding weather and COVID-19 and have important implications for understanding where the public stands with respect to current public health knowledge on COVID-19.

From January through June 2020, the weather’s impact on COVID-19 has been a present topic of discussion, where the volume of discussion ramped up between March 8 and April 1, coinciding with the beginning of stay-at-home orders throughout the United States. Furthermore, the spikes in the volume of discussion reflected significant events in the world. Figure 2 documents three such events: Trump’s comments in February claiming coronavirus would go away with the warm weather;^29^ the National Academies of Science’s response in early April to Trump’s February claims;^30^ and the White House’s promotion in late April of lab results suggesting heat slows coronavirus.^31^ This showed that Twitter conversation around the weather’s impact on the spread of COVID-19 correlated with an increase in the spread of the virus and, inferably, impacted individuals’ concerns.^15^

Figure 3 demonstrates a notable shift in opinion on the weather’s impact through the progression of the pandemic. While there was a significant decrease in tweets displaying uncertain opinions, there was an increase in the proportion of tweets claiming no effect and in those claiming some effect of the weather on the spread of COVID-19. Similarly, the non-trivial proportion of tweets identified by the Effect Classifier claiming some effect is noteworthy given that the scientific community has not reached a clear consensus of the weather’s impact on COVID-19.^30^ This claiming of an effect by users, regardless of whether it claims warming weather will improve or worsen the pandemic, shows that perception is formed as a result of broadcasted COVID-19 public health information and personal intuition on social media.

In Figure 4, where cluster topic frequencies are plotted over time, trends are shown in discussions about the weather’s impact on the spread of COVID-19. Furthermore, from January to February, there is a high frequency of discussion about cold weather and the flu, as these months exhibit both cold temperatures and the flu season, and the seasonality of COVID-19 was being discussed in reference to these topics. This was followed by an increase in discussion about reports made by scientific experts, from January 30 to March 19, about the weather’s impact on the spread of COVID-19, as the virus was just beginning to spread globally, and its seasonal behavior was unknown.

Simultaneously, there was an increase in discussion about Trump’s comments from February 13 to 27, on April 9 and after April 23, following the same pattern seen in Figure 2 where the three illustrated peaks occurred. The high frequency of the Trump cluster shows the impact of President’s statements and their constant relevance throughout the discussion of the weather’s impact on the spread of COVID- 19.

It is also interesting to note that the social distancing cluster did not show up in Figure 4 until April 2 and increased in frequency from May 7 to 28. This is likely because discussion about social distancing was not prevalent until after the nationwide lockdowns in the United States in late March, and the discussion increased as the weather got warmer and people were more tempted to avoid social distancing guidelines. Similarly, discussion about social distancing peaked the same day discussion about Trump peaked on April 23, when the White House promoted new evidence about heat possibly slowing the spread of COVID-19. This is curious, as many users claimed that heat will not slow the spread of COVID-19, only social distancing will.

Using clustering to reveal these topics helped understand which conversations were generating the greatest public response, allowing researchers to look into why these particular topics around the weather’s impact on COVID-19 were standing out. The clustering analysis revealed a structure to the data beyond the effect class framework that we pursued for the supervised learning. For instance, comparisons to the seasonality of influenza was a notable topic in size, yet sample tweets from that topic exhibited entirely different claims to the effect of weather (see S7). Therefore, our decision to include both our supervised and unsupervised analyses was verified by the different characteristics of the data revealed by each approach, which together enabled us to understand Twitter chatter.

During the manual annotation of tweets for effect, annotators recorded users’ proposed mechanisms for the impact of weather, which are of interest as they exhibit potential misconceptions or unfounded theories. Some users who expected warm weather to decrease coronavirus spread discussed the following mechanisms: sunlight increasing Vitamin D levels and boosting immune response to the virus; hot weather destroying the viral capsid; and higher malaria resistance in populations with warmer climates correlating with resistance to COVID-19. Conversely, some users believed that warm weather could negatively impact the pandemic due to an increased temptation to avoid social distancing guidelines, increased transmission through air conditioning units or higher humidity, and decreased compliance to wear recommended personal protective equipment. These mechanisms demonstrate that in the absence of consensus among experts, speculative theories can take hold on social media. Understanding the drivers of this information can inform public health response to the pandemic. From an NLP perspective, automatically detecting causal mechanisms from text could be integrated into opinion mining to summarize perceptions more quickly.^32^

This research is subject to limitations. As mentioned, the tri-class of “effect,” “no effect,” and “uncertain” problem proved too difficult for machine learning. Indeed, part of this arose from annotator difficulty in separating “no effect” and “uncertain” tweets. Several tweets were found to straddle the border of these two categories, partially due to the similarity of words across the “no effect” and “uncertain” tweets. This partly explains why collapsing these two categories into one improved our analysis performance enough to present results, and our adjusted Effect Classifier was able to successfully recognize users who claimed an effect.

An additional limitation in the effect annotation scheme was that we did not label for the magnitude of the effect. With this, we lose the nuance of whether tweets are claiming a strong, impactful or weak, inconsequential effect of the weather. One solution to this is to annotate for ‘weak’ or ‘strong’ effect or assign a numerical score for the strength of effect; with more ample training data it is plausible a model may successfully learn which tweets claim a strong effect or otherwise.

One significant language pattern that helped train our NLP analysis was the use of certain geographical locations to support a claim. For example, annotators noticed that warm locations, such as Florida and Singapore, were typically mentioned amongst users as a counterexample to undermine the possibility that warm weather will reduce the spread of COVID-19, and the names of these locations became a negative predictor for the “effect” class. Of course, not all mentions of warm locations in the data were as part of a counterexample, which exhibits one limitation of our model. Additionally, the Effect Classifier found the mention of “Trump” to be an accurate predictor for the “no effect/uncertain” class; this was largely due to sarcastic responses to Trump’s February predictions of the weather’s impact. Future directions include improving the performance of the Effect Classifier to detect more nuances of language, such as sarcasm and tone, which confused our models in some instances and are well-documented as difficult for machine learning models.^33^

## CONCLUSION

Our analyses revealed a surprising variety in conversations discussing potential seasonal impacts on COVID-19. The discussion went beyond the effect framework we chose that was centered around temperature and revealed broader beliefs on the impact of weather. For instance, the discussion around warm weather tempting the public to violate social distancing guidelines was unexpected, and points to an effect that has not yet been considered by researchers and could furthermore be modeled. Similarly, the presence of alternative facts such as increased air-conditioning use during warmer months worsening spread or increased transmission through mosquitos raises questions of how many subscribe to them. With these results in mind, social media can be used to crowdsource such mechanisms and provide topics for study in order to address public misconceptions. Especially during a pandemic, when everything is novel and unsettling for most, the understanding of public opinion is crucial for public health. In the future, computational methods could be used to detect public’s opinion in real-time from social media to prepare for pandemic responses. This study showed that not only is detecting public opinion on social media possible, but that careful attention should be placed on the individuality of perception and how to undermine misconceptions.

## Data Availability

All analysis processes are documented in the supplementary materials. Contact the corresponding author for the raw data.

## FUNDING

No funding was used to conduct this study.

## AUTHOR CONTRIBUTIONS

### Author Contributions

MG and BJ conducted pilot testing for data collection, and MG and AB worked jointly on final data collection, preparation, and machine learning classification analyses. JR, BJ, and MG annotated training data and contributed to qualitative analyses of data. MG designed the topic analysis, for which BJ, AO, AB, and MG wrote code and BJ executed. AO conducted validation testing reported in the supplementary materials. JR led the drafting of the manuscript with assistance from BJ, MG, AB, and MSJ. MSJ conceived the study, supervised the project, and revised the manuscript for important intellectual content.

## ACKNOWLEDGMENTS

We thank Yicheng Wang and Elizabeth Mason who provided feedback and suggestions on earlier versions of this manuscript. We also thank Catherine DiGennaro for her contribution in framing the research project.

## CONFLICT OF INTEREST STATEMENT

None declared.

## Supplementary Material for

### S1. Tweet Collection

#### S1.1. Query Decision

We tested queries consisting of two parts: a term describing COVID-19 (‘coronavirus,’ ‘covid,’ or ‘covid19’) and a combination of weather keywords (‘weather’, ‘temperature’, ‘climate’, ‘humidity’). Queries were directly tested on Twitter’s website. The inclusion of ‘weather’ as a search term was necessary given the topic, and upon manual inspection, we determined that the queries with keywords ‘temperature’ and ‘climate’ returned a relatively low volume of tweets with largely irrelevant results.

Results including ‘temperature’ tended to focus on symptoms associated with coronavirus infection, such as fevers and chills. Results including ‘climate’ tended to focus on discussion regarding the intersection between COVID-19 and climate change. Results for ‘humidity’ were often related but had low volumes. We decided not to include any additional terms due to the low volumes and to reduce the amount of filtering.

We made use of the Twitter search operators (see https://developer.twitter.com/en/docs/tweets/search/guides/premium-operators) to return tweets in English, and only original tweets, not retweets. The operators for doing so were “-is:retweet” and “lang:en,” which are included in the query.

#### S1.2. Counts Endpoint

Twitter provides a Counts Endpoint under their premium tier of service to return the number of tweets matching a query over a given time span (see https://developer.twitter.com/en/docs/tweets/search/api-reference/premium-search). This number is an upper bound, since the count may include deleted tweets that will not be returned with an actual query. The Counts Endpoint returned a count of 174,987 for the query “(coronavirus OR covid OR covid19) weather -is:retweet lang:en” between January 23, 2020 and June 22, 2020.

#### S1.3. Article and Quoted/Replied-to Tweet Collection

For tweets sharing an article or quoting/replying to another tweet, if available, we gathered the article headline and description and the quoted/replied-to tweet text, to factor in with the analyses. Twitter provides some of this info in the returned tweet object – the article headline/description, and quoted tweet object are included, as well as the ID of the replied-to tweet. Using Twitter’s API, we collected all such replied-to tweets and linked them to their referencing tweet.

A small number of fetched tweets contained URL’s of news articles but did not have any attached news article data. To ensure we had all possible data, we visited each of these URL’s and extracted from the website’s HTML the headline/description. Websites containing this information will include a tag for Twitter in their HTML, which we searched for to ensure that we collected the article data as presented on Twitter. Code for this collection is available on the Github.

### S2. Preprocessing

Before feeding data into any of these steps, we first preprocessed tweets with the following five steps:

1) Removing any HTML, non-ASCII text, and emojis from the tweet text.
2) Removing any references to popular weather channels, which were picked up by our query and generally false positives.
3) Removing any “mentions,” where a user tags another user, from tweets. The exception to this was President Trump’s Twitter account, @realDonaldTrump, was replaced with “Trump”;
4) Removing any trailing hashtags, where trailing hashtags are a chain of hashtags present at the end of a tweet meant to associate the tweet with a topic. Any hashtags within the middle of the tweet were kept, with the hashtag symbol removed.
5) Tokens were then standardized to lower-case form and standardized further with lemmatization (mapping each word to its root form, e.g., “running” to “run”) and stemming (removing suffixes from words, e.g., “chairs” to “chair”).

For more details see the Github repository.

The decision to replace Trump’s Twitter handle with his name was to provide additional context for tweets referring to him. As shown in the clustering results, Trump’s February comments on weather’s impact drove a substantial amount of discussion on Twitter about the weather’s potential impacts, with several tweets referring to him by name or by his account handle. Given his status as a public figure and the lingering discussion around his comments, we standardized referenced to him to one form in order to better track discussion relating to him.

### S3. Rule-Based Filtering and Rule Performance

#### S3.1. Detailed Rule Descriptions

As mentioned in the article, we employed rule-based filtering to narrow our corpus down and remove noise. The rules-based filtering consisted of three rules applied sequentially, which discards a tweet if it fails the rule. Figure S1 gives a high-level overview of the mechanics of each rule, and comments on the rules follow.

**Figure S1:**
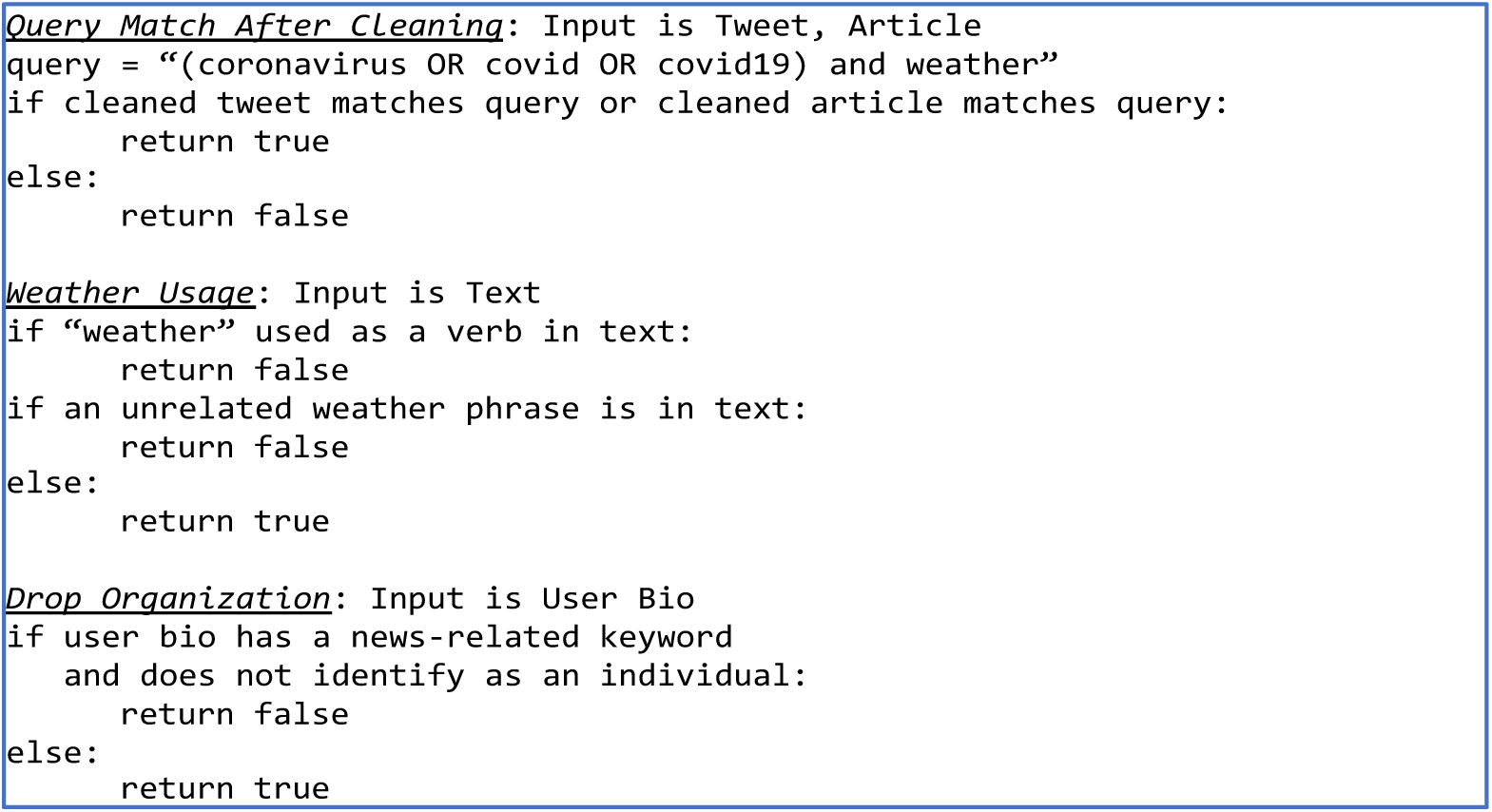
Logic for tree Rule-Based Filters.

Rules are applied sequentially. In this context, “true” means we kept the tweet and “false” means we discarded it. A “cleaned tweet/article” is one that has been processed as described in S2.

##### Query Match After Cleaning

This rule filtered out false positives due to the sheer popularity of our keywords (e.g., a tweet may comment on pleasant weather and end with “#coronavirus), and removed tweets where the keywords were split across different parts of the tweet (e.g., “weather” appeared in the article text, and “covid” in the tweet itself), as such tweets were generally found to be irrelevant. This rule dropped 32,054 tweets.

##### Weather Usage

This rule assessed tweet relevancy by limiting the dataset to tweets using the desired form of the keyword “weather” as a noun, not as a verb or in an idiom (e.g. “under the weather”). This rule dropped 42,387 tweets.

##### Posted by Organization

This rule restricted the tweets to those posted by individuals, who are the focus of study, not news organizations. It detected organizations by checking if the user’s bio contained a news-organization keyword, or by seeing if the bio mentioned an individual-occupation keyword (e.g. “reporter”) or first-person pronoun. This rule dropped 7,372 tweets.

#### S3.2. Rule Performance

To assess our rule-based filters, a sample of 200 tweets discarded at each step were inspected to look for false negatives. Here, false negative are tweets that were truly relevant but that were discarded by the rule.

The “Query Match After Cleaning” filter had 16 false negatives (false negative rate= 16/200 = 8%). The “Weather Usage” filter had 13 false negatives (false negative rate= 13/200 = 6.5%). The “Drop Organization” filter had 12 false negatives (false negative rate= 12/200 = 6%). The low false negative rates for all these filters suggest that the filters did not discard a significant amount of relevant tweets.

#### S3.3. Attempted Rule Based Relevancy Classification

The high performance of all classifiers for the Relevancy Classification raises the question of whether the problem is too simple for machine learning, and if it could be replaced by simple rule-based logic. To explore this, a rule-based classifier was written after manual inspection and looking at the various keywords with their respective weights that the ML algorithm produced.

In our simple rule-based relevancy classifier, a tweet was classified as relevant if it contained, after lemmatizing and stemming, any of the following words: (‘kill’, ‘outbreak’, ‘away’, ‘scientific’, ‘study’, ‘distance’, ‘death’, ‘report’, ‘help’, ‘slow’, ‘curb’, ‘reduce’, ‘increase’), and none of the following: (‘plan’, ‘countries’, ‘state’, ‘home’, ‘forecast’, ‘closed’, ‘pleasant’, ‘beautiful’, ‘nice’, ‘beaches’), which were generally found to be in unrelated tweets.

Table S1 shows a confusion matrix for the attempted rule-based classifier. The overall rule-based accuracy was 0.67, suggesting that our manual rules were not good enough to replace machine learning. Indeed, if there were a simple rule-based method for the problem a decision tree likely would have discovered it.[1]

**Table S1:** Confusion Matrix, showing in absolute counts true positives (TP), false negatives (FN), false positives (FP), and true negatives (TN), from rule-based filters. Total number of samples is 2,786.

| Actual \ Predicted | Unrelated | Related |
| --- | --- | --- |
| Unrelated | 1,362 (TN) | 564 (FP) |
| Related | 368 (FN) | 492 (TP) |

### S4. Supervised Learning Design Decisions

#### S4.1. Concatenating Tweet, Article, and Reply for Relevancy and Effect Classification

As mentioned in the article, for each tweet collected, the tweet text, article headline and description (if any), and quoted or replied-to tweet (if any) were merged into one text sample to be analyzed be our classifiers. The text in addition to the tweet text, which includes any article text, quoted or replied-to-tweet text, are referred to as our “reference” text as this additional text provides necessary context to the tweet text. Below we further describe the motivation for this reference text.

The reason for including referenced texts in the Relevancy Classifier was to provide additional context for seeing if the tweet is related to our study. The inclusion aided annotators in deciding the class label, and so the same information was supplied to the Relevancy Classifier.

The referenced texts were also included for the Effect Classifier to account for tweets that endorsed the opinion stated in the referenced text (such as tweets that share and comment on a news article). One potential issue that was discussed was disagreements between the user’s tweet and the referenced text, but during annotation the Effect Class of the user’s text and the referenced text rarely differed. Due to this and out of simplicity (i.e. to avoid having to predict the opinion of each text part and then aggregate them) we predicted the classification of “effect” vs. “no effect/uncertain” based on all text.

For the type and direction of effect (e.g. “improve with warmer weather”), the similar issue of disagreement between the user and referenced text also arose. To handle this, annotators recorded the type and direction of effect based on the user’s opinion, which was separated from the referenced text. Because we did not attempt machine learning on the type and direction of effect, the issue of what data to supply the model never arose.

#### S4.2. Text Featurization: Other Featurization Methods

As mentioned in the article, three featurization methods – Bag of Words (BOW), Term Frequency-Inverse Document Frequency (TF-IDF), and Embeddings from Language Models (ELMO) – were used to generate vectorized inputs for the machine learning models. A fourth featurization method, Word2Vec, was briefly explored. Word2Vec is a word-embedding model that maps each word in a text corpus to a vector in some high-dimensional space, where the representation of the word depends on the context surrounding it.[2] Once a representation is built for each word in a text corpus, a document (tweet) can be represented as the average of its word’s vectors. While Word2Vec seemed promising due to its ability to factor in context, preliminary results showed that Word2Vec’s performance was subpar, potentially due to the short, limited context provided by tweets. Therefore, we did not formally include it in our analysis.

#### S4.3. Supervised Learning (Relevancy and Effect) Training Workflow

Figure S2 shows a schematic for the training process for the two classifiers used. For our binary classifications, we optimized with respect to average precision (ROC-PR); for the trinary classification (“effect” vs “no effect” vs “uncertain”), we optimized with respect to balanced accuracy. After a first round of inner cross-validation, the top features are selected from one of the best initial classifiers, Gradient Descent SVM. Top features were determined by their coefficients for the initial Gradient Descent SVM classifier and were taken if their coefficient is above 1e-5 in absolute value.

**Figure S2:**
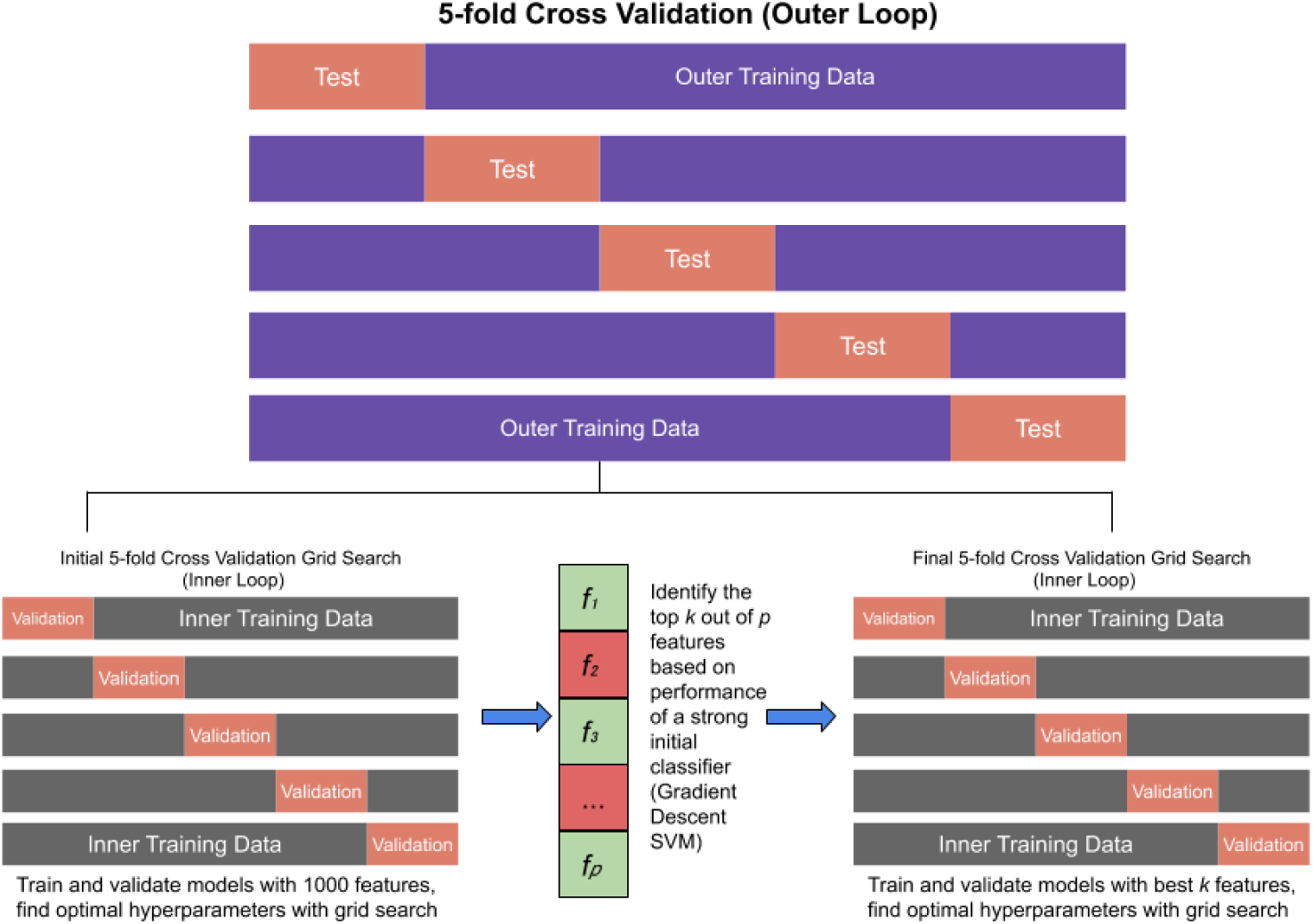
Supervised Learning Training Workflow.

#### S4.4. Supervised Learning Hyperparameters

The machine learning hyperparameters are listed below in Table S2.

**Table S2:** Machine Learning Hyperparameters

| Model Name | Parameter | Values Explored |
| --- | --- | --- |
| Ridge Classifier | Alpha | 0.01, 1.0 |
| KNN | Number of Neighbors | 5, 10, 15 |
|  | Leaf Size | 20, 30, 40 |
|  | Weights | Uniform, Distance |
| Random Forest Classifier | Maximum Depth | 2, 3, 4, 5, None |
|  | CCP Alpha | 0.0, 0.01, 0.1, 0.5 |
|  | Use out of bag samples (oob_score) | True, False |
| Linear SVM | Regularization parameter (C) | 0.5, 1, 2, 5, 10, 100, 500, 1000 |
|  | Loss | Hinge, Squared Hinge |
|  | Penalty | L1, L2 |
| Gradient Descent SVM | Regularization multiplier (Alpha) | 0.0001, 0.0005, 0.001, 0.01, 0.1, 1 |
| Gradient Descent LR | Regularization multiplier (Alpha) | 0.0001, 0.0005, 0.001, 0.01, 0.1, 1 |
| Logistic Regression | Inverse of Regularization strength | 0.5, 1.0, 2, 5, 10, 100 |
| Naïve Bayes Classifiers | Additive smoothing parameter (Alpha) | 0.01, 0.05, 0.1, 1.0, 2.0 |
| Decision Tree | Maximum Depth | 2, 3, 4, 5, None |
|  | CCP Alpha | 0.0, 0.01, 0.1, 0.5 |

### S5. Detailed Classifier Performances

#### S5.1. Relevancy Classifier Metrics

The relevancy classifier metrics are listed below in Table S3.

**Table S3:**
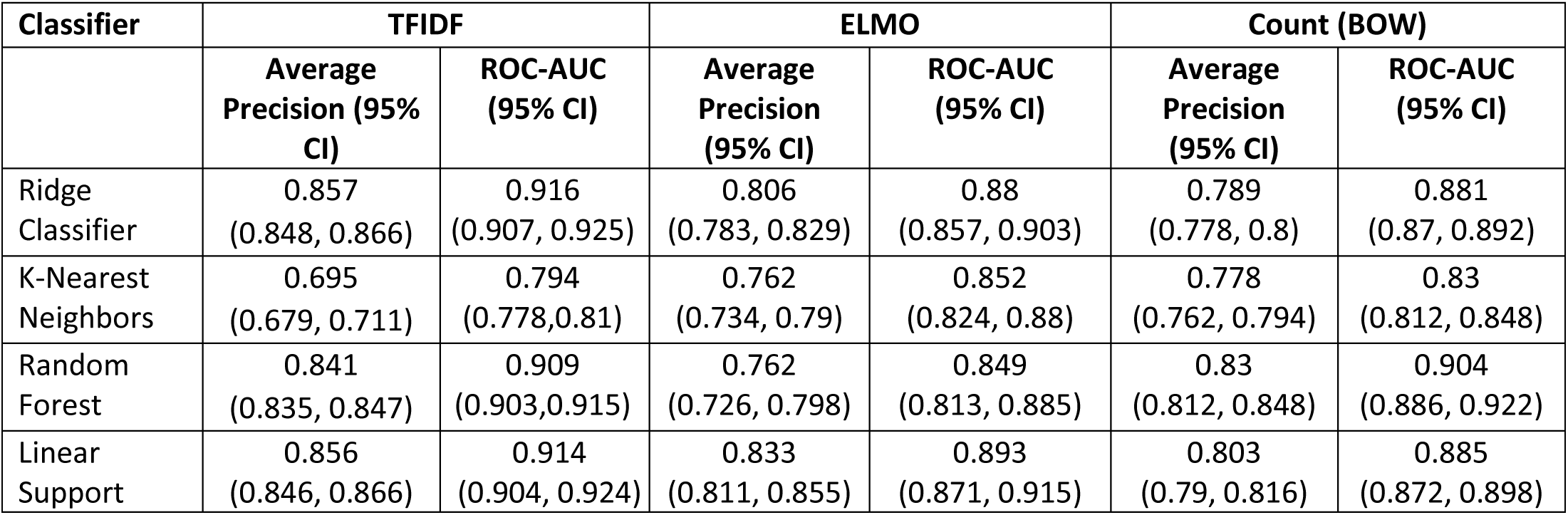
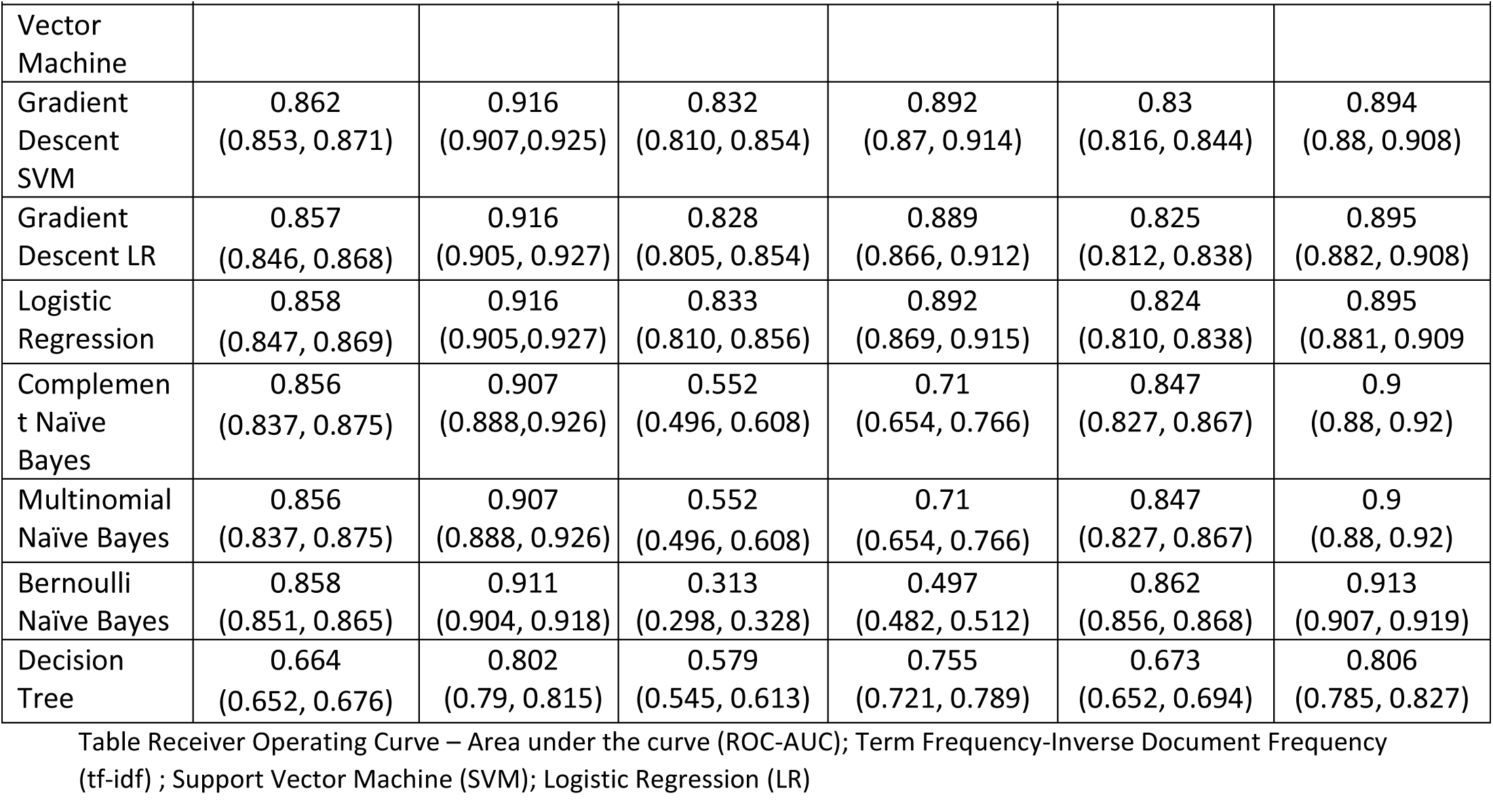
Relevancy Classifier Metrics

#### S5.2. Effect Classifier Metrics

##### S5.2.1. Three-way Effect Classification

The three-way effect classifier metrics are listed in Table S4 below. These metrics are the results of the effect classifier with the effect scheme “effect”, “no effect” and “uncertain.”

**Table S4:** Three-Way Effect Classifier Metrics

| Model Name | Count (BOW) | ELMO | TFIDF |
| --- | --- | --- | --- |
|  | **Balanced Accuracy (95% CI)** |  |  |
| Ridge Classifier | 0.486<br>(0.455, 0.517) | 0.516<br>(0.487, 0.545) | 0.517<br>(0.484, 0.55) |
| KNN | 0.435<br>(0.392, 0.478) | 0.45<br>(0.421, 0.479) | 0.448<br>(0.419, 0.477) |
| Random Forest Classifier | 0.508<br>(0.477, 0.539) | 0.482<br>(0.477, 0.487) | 0.503<br>(0.475, 0.531) |
| Linear SVM | 0.486<br>(0.452, 0.52) | 0.526<br>(0.498, 0.554) | 0.514<br>(0.479, 0.549) |
| Gradient Descent SVM | 0.502<br>(0.47, 0.534) | 0.484<br>(0.447, 0.521) | 0.514<br>(0.481, 0.547) |
| Gradient Descent LR | 0.505<br>(0.471, 0.539) | 0.493<br>(0.45, 0.536) | 0.511<br>(0.481, 0.541) |
| Logistic Regression | 0.497<br>(0.465, 0.529) | 0.52<br>(0.488, 0.552) | 0.503<br>(0.476, 0.53) |
| Multinomial Naïve Bayes | 0.502<br>(0.469, 0.535) | 0.421<br>(0.39, 0.452) | 0.491<br>(0.455, 0.527) |
| Complement Naïve Bayes | 0.506<br>(0.462, 0.55) | 0.426<br>(0.404, 0.448) | 0.515<br>(0.479, 0.551) |
| Bernoulli Naïve Bayes | 0.5<br>(0.47, 0.53) | 0.34<br>(0.334, 0.346) | 0.495<br>(0.465, 0.524) |
| Decision Tree | 0.411<br>(0.395, 0.426) | 0.338<br>(0.362, 0.414) | 0.433<br>(0.41, 0.456) |

Confidence Interval (CI);

##### S5.2.2. Effect vs The Rest

The two-way effect classifier metrics are listed below in Table S5. These metrics are the results of the effect classifier after grouping together the “uncertain” and “no effect” categories, to have a classifier that categorized tweets as either “effect” or “the rest.”

**Table S5:** Effect Classifier Metrics using groups “effect” vs “the rest”

| Classifier | TFIDF |  | ELMO |  | Count (BOW) |  |
| --- | --- | --- | --- | --- | --- | --- |
|  | Average Precision (95% CI) | ROC-AUC (95% CI) | Average Precision (95% CI) | ROC-AUC (95% CI) | Average Precision (95% CI) | ROC-AUC (95% CI) |
| Ridge Classifier | 0.551<br>(0.532, 0.570) | 0.762<br>(0.743, 0.781) | 0.54<br>(0.522, 0.558) | 0.762<br>(0.744, 0.78) | 0.498<br>(0.467, 0.529) | 0.717<br>(0.686, 0.748) |
| K-Nearest Neighbors | 0.468<br>(0.424, 0.512) | 0.692<br>(0.648, 0.736) | 0.47<br>(0.433, 0.507) | 0.702<br>(0.665, 0.739) | 0.467<br>(0.437, 0.497) | 0.707<br>(0.677, 0.737) |
| Random Forest | 0.502<br>(0.469, 0.535) | 0.724<br>(0.691, 0.757) | 0.479<br>(0.445, 0.513) | 0.716<br>(0.682, 0.75) | 0.521<br>(0.499, 0.543) | 0.761<br>(0.739, 0.783) |
| Linear Support Vector Machine | 0.544<br>(0.52, 0.568) | 0.759<br>(0.735, 0.783) | 0.543<br>(0.523, 0.563) | 0.759<br>(0.739, 0.779) | 0.518<br>(0.488, 0.548) | 0.722<br>(0.692, 0.752) |
| Gradient Descent SVM | 0.561<br>(0.542, 0.58) | 0.768<br>(0.749, 0.787) | 0.56<br>(0.543, 0.577) | 0.77<br>(0.753, 0.787) | 0.529<br>(0.501, 0.557) | 0.734<br>(0.706, 0.762) |
| Gradient Descent LR | 0.55<br>(0.531, 0.569) | 0.762<br>(0.743, 0.781) | 0.554<br>(0.539, 0.569) | 0.771<br>(0.756, 0.786) | 0.549<br>(0.525, 0.573) | 0.745<br>(0.721, 0.769) |
| Logistic Regression | 0.544<br>(0.524, 0.564) | 0.758<br>(0.738, 0.778) | 0.556<br>(0.541, 0.571) | 0.768<br>(0.753, 0.783) | 0.545<br>(0.52, 0.57) | 0.743<br>(0.718, 0.768) |
| Complement Naïve Bayes | 0.556<br>(0.524, 0.588) | 0.768<br>(0.736, 0.8) | 0.407<br>(0.37, 0.444) | 0.673<br>(0.636, 0.710) | 0.534<br>(0.504, 0.564) | 0.753<br>(0.723, 0.783) |
| Multinomial Naïve Bayes | 0.556<br>(0.524, 0.564) | 0.768<br>(0.736, 0.8) | 0.407<br>(0.37, 0.444) | 0.673<br>(0.636, 0.710) | 0.534<br>(0.504, 0.564) | 0.753<br>(0.723, 0.783) |
| Bernoulli Naïve Bayes | 0.537<br>(0.508, 0.566) | 0.753<br>(0.724, 0.782) | 0.278<br>(0.269, 0.287) | 0.511<br>(0.502, 0.52) | 0.529<br>(0.5, 0.558) | 0.749<br>(0.72, 0.778) |
| Decision Tree | 0.363<br>(0.327, 0.399) | 0.647<br>(0.611, 0.683) | 0.367<br>(0.332, 0.402) | 0.629<br>(0.594, 0.664) | 0.373<br>(0.339, 0.407) | 0.662<br>(0.628, 0.696) |

Receiver Operating Curve – Area under the curve (ROC-AUC); Term Frequency-Inverse Document Frequency (tf-idf); Support Vector Machine (SVM); Logistic Regression (LR); Confidence Interval (CI)

### S6. Exploratory Analysis: Top 10 Dates by Volume with Most Frequent News Headline

Based on the day-wise distribution of the Tweets, we found the ten days with most tweets and reported the most discussed news article from that day. These headlines with the corresponding days are shown below in Table S6.

**Table S6:**
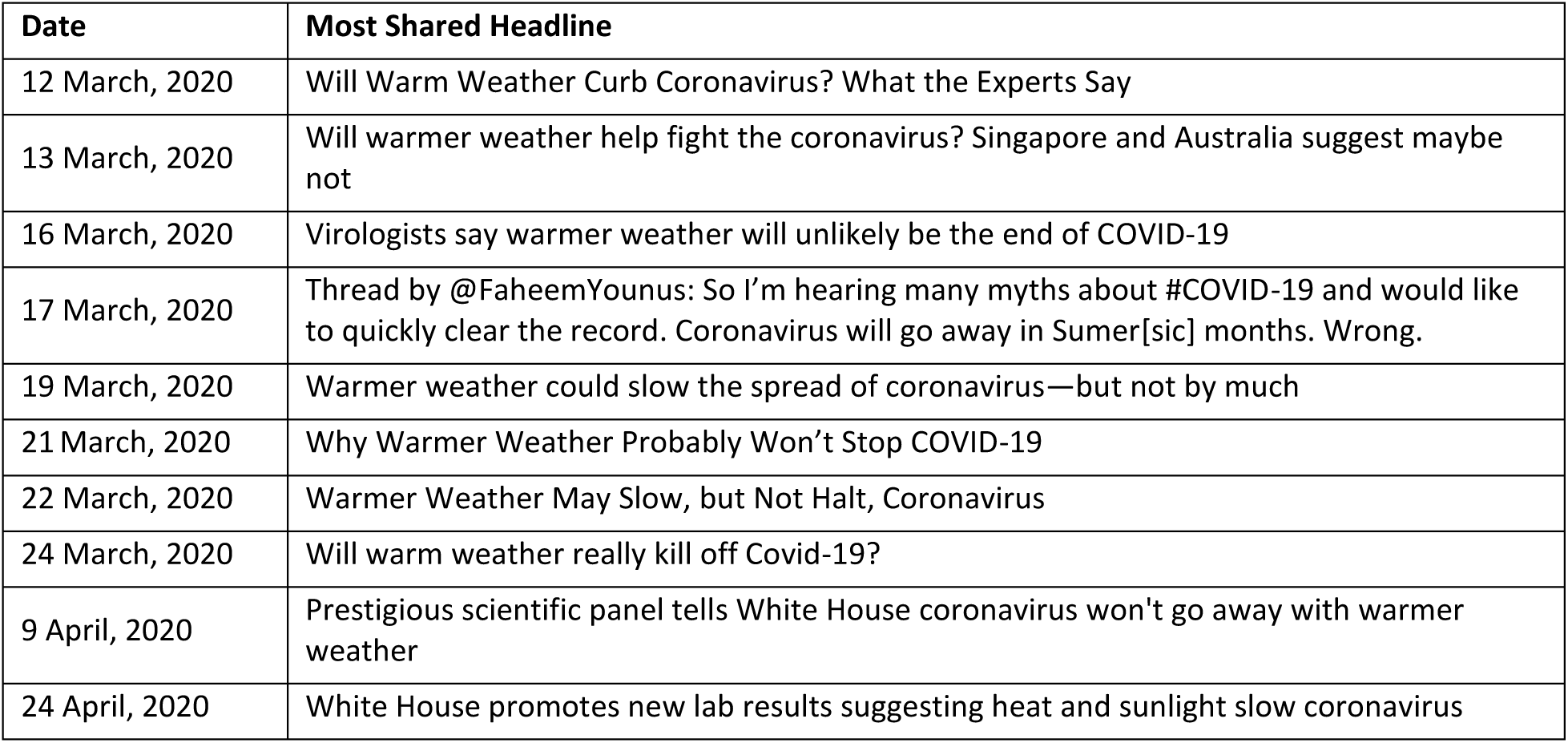
Top 10 dates with highest tweet volume and the corresponding most shared headline that day, sorted by date

| Date | Most Shared Headline |
| --- | --- |
| 12 March, 2020 | Will Warm Weather Curb Coronavirus? What the Experts Say |
| 13 March, 2020 | Will warmer weather help fight the coronavirus? Singapore and Australia suggest maybe not |
| 16 March, 2020 | Virologists say warmer weather will unlikely be the end of COVID-19 |
| 17 March, 2020 | Thread by @FaheemYounus: So I'm hearing many myths about #COVID-19 and would like to quickly clear the record. Coronavirus will go away in Sumer[sic] months. Wrong. |
| 19 March, 2020 | Warmer weather could slow the spread of coronavirus—but not by much |
| 21 March, 2020 | Why Warmer Weather Probably Won't Stop COVID-19 |
| 22 March, 2020 | Warmer Weather May Slow, but Not Halt, Coronavirus |
| 24 March, 2020 | Will warm weather really kill off Covid-19? |
| 9 April, 2020 | Prestigious scientific panel tells White House coronavirus won't go away with warmer weather |
| 24 April, 2020 | White House promotes new lab results suggesting heat and sunlight slow coronavirus |

In Table S6, where the top ten days with the highest volume of tweets were pulled, it is clear most of the dates with the highest volume of tweets occurred in the time period from March 13^th^ to March 22^nd^ when there was an overall increase in twitter activity. Therefore, to isolate key events (and not just pull the top ten days by volume) and identify the dates where twitter activity significantly increased, we identified the top ten local maxima (i.e., peaks) in the tweet volumes. After identifying these local maxima, we pulled the most shared headline from each day where the maximum occurred. Out of the ten local maxima, three key peaks were presented in Figure 2 in the article. These three key peaks were the only maxima that corresponded to real world events. The top ten local maxima and the corresponding headlines are shown in Table S7 below.

**Table S7:** Top 10 dates where a local maximum occurred for tweet volume and the most shared headline that day, sorted by date

| Date | Most Shared Headline |
| --- | --- |
| *11 February, 2020 | Trump says Coronavirus will be gone by April when the weather gets warmer, doesn't offer scientific explanation |
| 26 February, 2020 | Can Coronavirus Be Crushed By Warmer Weather? |
| 13 March, 2020 | Will warmer weather help fight the coronavirus? Singapore and Australia suggest maybe not |
| 17 March, 2020 | Thread by @FaheemYounus: So I'm hearing many myths about #COVID-19 and would like to quickly clear the record. Coronavirus will go away in Sumer months. Wrong. |
| 19 March, 2020 | Warmer weather could slow the spread of coronavirus—but not by much |
| 21 March, 2020 | Why Warmer Weather Probably Won't Stop COVID-19 |
| 24 March, 2020 | Will warm weather really kill off Covid-19? |
| *9 April, 2020 | Prestigious scientific panel tells White House coronavirus won't go away with warmer weather |
| *24 April, 2020 | White House promotes new lab results suggesting heat and sunlight slow coronavirus |

\*Refers to one of the three key peaks

### S7. Clustering

We tested *k* = 10, 15, 20, 25, and 30 on three clustering algorithms: *k*-means, *k*-medoids, and latent Dirichlet allocation (LDA). We used term-frequency inverse document-frequency weighting over ELMo contextual vectors because the former approach retains features to a word map, allowing them to be more interpretable.[3] The decision to choose *k* = 25 for the clustering algorithm was determined primarily by a qualitative approach. Under the *k*-means clustering, qualitative analysis showed that the topic clusters were not well separable at *k* < 25 or *k* > 25. Specifically, with *k* < 25, every cluster had similar volume and largely consisted of generic tweets about the influence of weather on coronavirus spread. With *k* > 25, one or two clusters dominated, and these clusters were comprised of generic tweets, while the other clusters, although less generic, maintained a low volume. High *k* values tended to reflect noise in the data, especially phrases used commonly in tweets that did not provide any common topic linking the clusters together. Thus, we did not test beyond *k* = 30. At *k* = 25, we deduced important topics (e.g., social distancing, Trump administration, influenza comparisons) that maintained a significant volume (see Table S8).

The k-medoids algorithm failed to provide separation at any value of *k*; we speculate that this is because the method anchors a cluster’s centroid on one of the tweets, creating several low volume clusters with repeated tweets and one high volume cluster with generic tweets.

The LDA algorithm failed to provide separation at any value of *k*; we speculate that this is because the corpus was already significantly narrowed during the relevancy classification phase. Thus, it may be difficult for LDA to distinguish among a corpus discussing relatively similar content.

These conclusions are supported by Table S9, which highlights each algorithm’s ability to separate topics of discussion into distinct clusters. It is clear that the *k*-means algorithm appropriately partitions text throughout all clusters, while *k*-medoids and LDA partition the majority of text to one cluster.

**Table S8:**
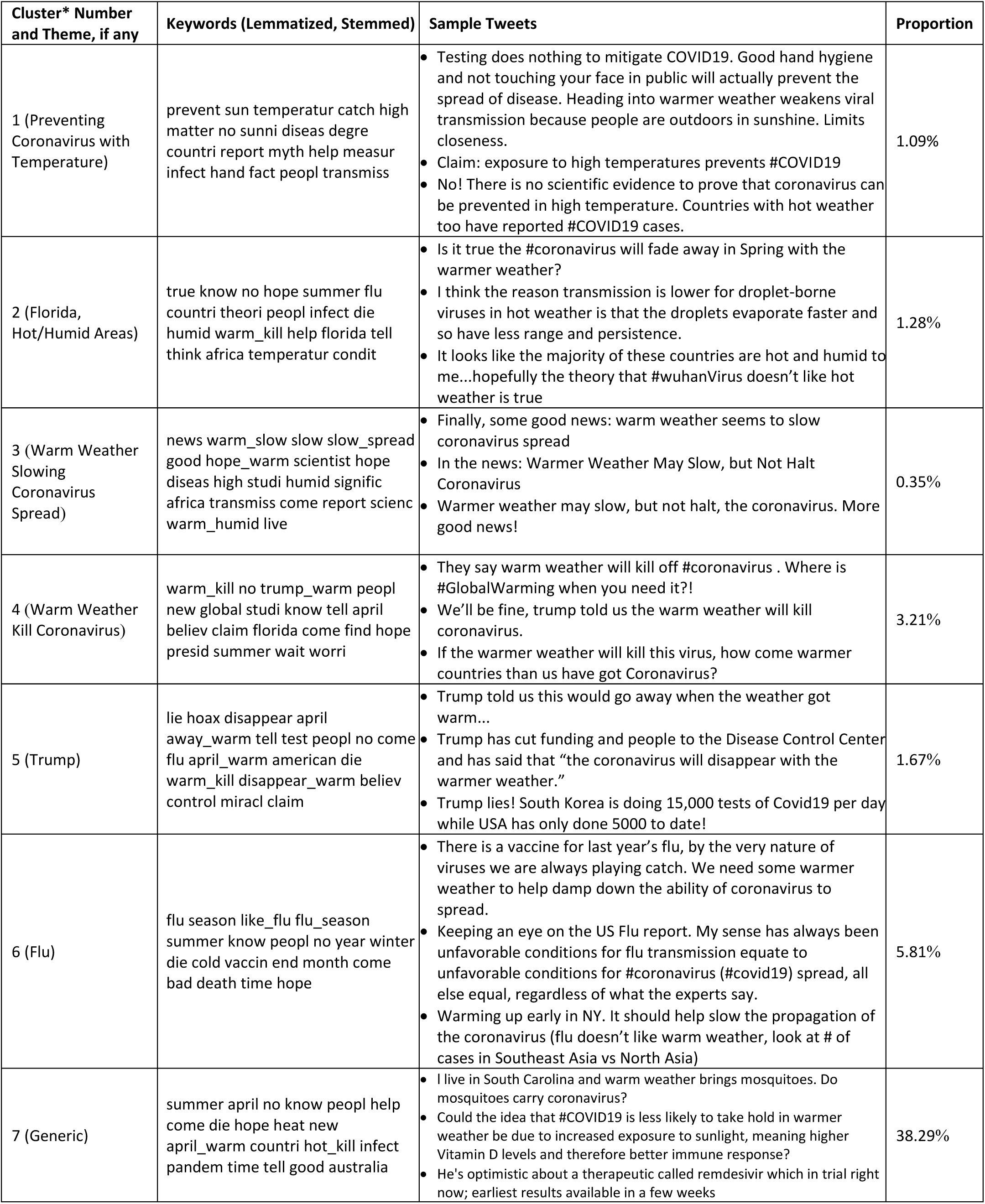
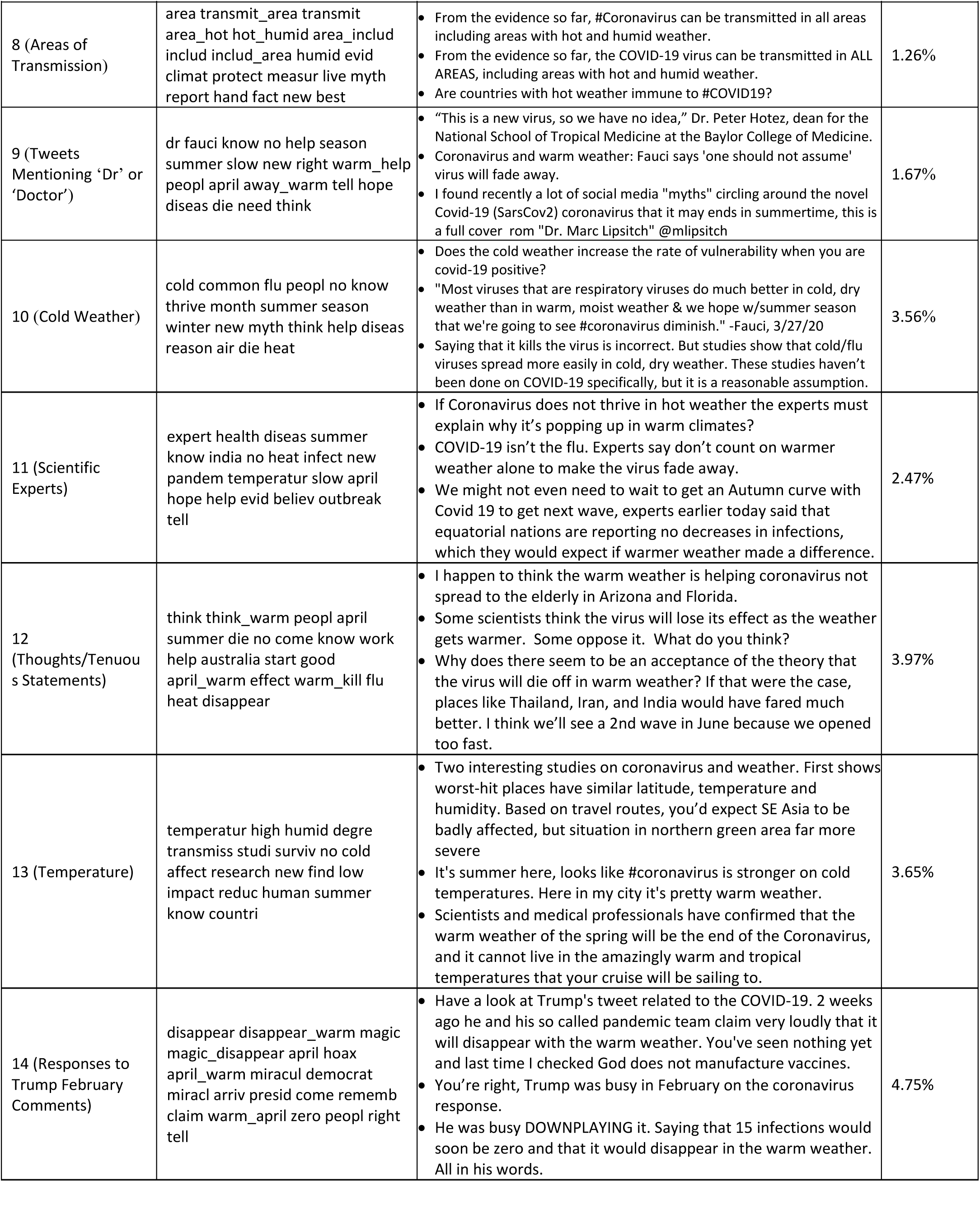
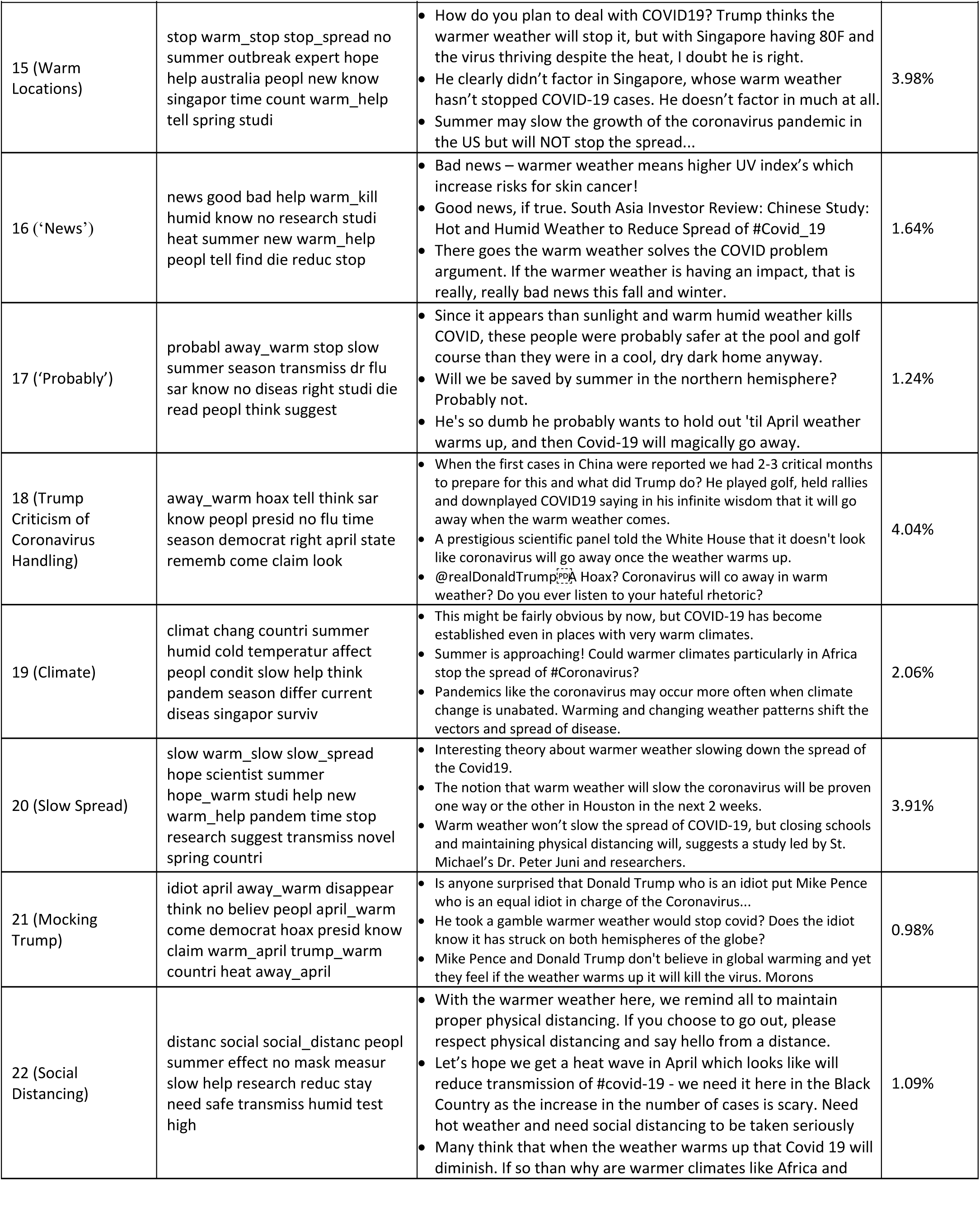
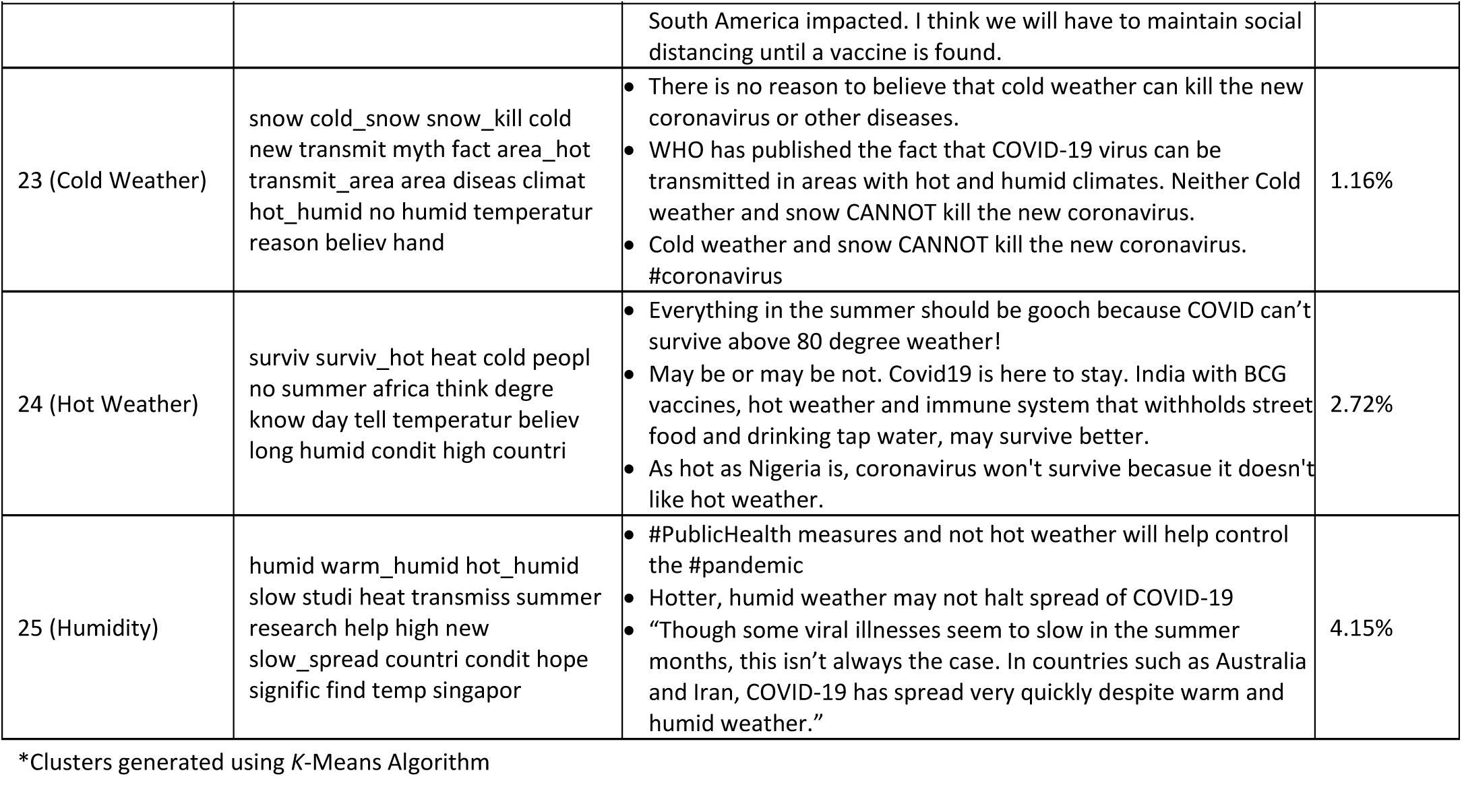
Top Key Words for the 25 Clusters with Sample Tweets

| Cluster* Number and Theme, if any | Keywords (Lemmatized, Stemmed) | Sample Tweets | Proportion |
| --- | --- | --- | --- |
| 1 (Preventing Coronavirus with Temperature) | prevent sun temperatur catch high matter no sunni diseas degre countri report myth help measur infect hand fact peopl transmiss | Testing does nothing to mitigate COVID19. Good hand hygiene and not touching your face in public will actually prevent the spread of disease. Heading into warmer weather weakens viral transmission because people are outdoors in sunshine. Limits closeness.<br>Claim: exposure to high temperatures prevents #COVID19<br>No! There is no scientific evidence to prove that coronavirus can be prevented in high temperature. Countries with hot weather too have reported #COVID19 cases. | 1.09% |
| 2 (Florida, Hot/Humid Areas) | true know no hope summer flu countri theori peopl infect die humid warm_kill help florida tell think africa temperatur condit | Is it true the #coronavirus will fade away in Spring with the warmer weather?<br>I think the reason transmission is lower for droplet-borne viruses in hot weather is that the droplets evaporate faster and so have less range and persistence.<br>It looks like the majority of these countries are hot and humid to me...hopefully the theory that #wuhanVirus doesn't like hot weather is true | 1.28% |
| 3 (Warm Weather Slowing Coronavirus Spread) | news warm_slow slow slow_spread good hope_warm scientist hope diseas high studi humid signific africa transmiss come report scienc warm_humid live | Finally, some good news: warm weather seems to slow coronavirus spread<br>In the news: Warmer Weather May Slow, but Not Halt Coronavirus<br>Warmer weather may slow, but not halt, the coronavirus. More good news! | 0.35% |
| 4 (Warm Weather Kill Coronavirus) | warm_kill no trump_warm peopl new global studi know tell april believ claim florida come find hope presid summer wait worri | They say warm weather will kill off #coronavirus . Where is #GlobalWarming when you need it?!<br>We'll be fine, trump told us the warm weather will kill coronavirus.<br>If the warmer weather will kill this virus, how come warmer countries than us have got Coronavirus? | 3.21% |
| 5 (Trump) | lie hoax disappear april away_warm tell test peopl no come flu april_warm american die warm_kill disappear_warm believ control miracl claim | Trump told us this would go away when the weather got warm...<br>Trump has cut funding and people to the Disease Control Center and has said that "the coronavirus will disappear with the warmer weather."<br>Trump lies! South Korea is doing 15,000 tests of Covid19 per day while USA has only done 5000 to date! | 1.67% |
| 6 (Flu) | flu season like_flu flu_season summer know peopl no year winter die cold vaccin end month come bad death time hope | There is a vaccine for last year's flu, by the very nature of viruses we are always playing catch. We need some warmer weather to help damp down the ability of coronavirus to spread.<br>Keeping an eye on the US Flu report. My sense has always been unfavorable conditions for flu transmission equate to unfavorable conditions for #coronavirus (#covid19) spread, all else equal, regardless of what the experts say.<br>Warming up early in NY. It should help slow the propagation of the coronavirus (flu doesn't like warm weather, look at # of cases in Southeast Asia vs North Asia) | 5.81% |
| 7 (Generic) | summer april no know peopl help come die hope heat new april_warm countri hot_kill infect pandem time tell good australia | I live in South Carolina and warm weather brings mosquitoes. Do mosquitoes carry coronavirus?<br>Could the idea that #COVID19 is less likely to take hold in warmer weather be due to increased exposure to sunlight, meaning higher Vitamin D levels and therefore better immune response?<br>He's optimistic about a therapeutic called remdesivir which in trial right now; earliest results available in a few weeks | 38.29% |
| 8 (Areas of Transmission) | area transmit_area transmit<br>area_hot hot_humid area_includ<br>includ includ_area humid evid<br>climat protect measur live myth<br>report hand fact new best | From the evidence so far, #Coronavirus can be transmitted in all areas including areas with hot and humid weather.<br>From the evidence so far, the COVID-19 virus can be transmitted in ALL AREAS, including areas with hot and humid weather.<br>Are countries with hot weather immune to #COVID19? | 1.26% |
| 9 (Tweets Mentioning 'Dr' or 'Doctor') | dr fauci know no help season<br>summer slow new right warm_help<br>peopl april away_warm tell hope<br>diseas die need think | "This is a new virus, so we have no idea," Dr. Peter Hotez, dean for the National School of Tropical Medicine at the Baylor College of Medicine.<br>Coronavirus and warm weather: Fauci says 'one should not assume' virus will fade away.<br>I found recently a lot of social media "myths" circling around the novel Covid-19 (SarsCov2) coronavirus that it may ends in summertime, this is a full cover rom "Dr. Marc Lipsitch" @mlipsitch | 1.67% |
| 10 (Cold Weather) | cold common flu peopl no know<br>thrive month summer season<br>winter new myth think help diseas<br>reason air die heat | Does the cold weather increase the rate of vulnerability when you are covid-19 positive?<br>"Most viruses that are respiratory viruses do much better in cold, dry weather than in warm, moist weather & we hope w/summer season that we're going to see #coronavirus diminish." -Fauci, 3/27/20<br>Saying that it kills the virus is incorrect. But studies show that cold/flu viruses spread more easily in cold, dry weather. These studies haven't been done on COVID-19 specifically, but it is a reasonable assumption. | 3.56% |
| 11 (Scientific Experts) | expert health diseas summer<br>know india no heat infect new<br>pandem temperatur slow april<br>hope help evid believ outbreak<br>tell | If Coronavirus does not thrive in hot weather the experts must explain why it's popping up in warm climates?<br>COVID-19 isn't the flu. Experts say don't count on warmer weather alone to make the virus fade away.<br>We might not even need to wait to get an Autumn curve with Covid 19 to get next wave, experts earlier today said that equatorial nations are reporting no decreases in infections, which they would expect if warmer weather made a difference. | 2.47% |
| 12 (Thoughts/Tenuous Statements) | think think_warm peopl april<br>summer die no come know work<br>help australia start good<br>april_warm effect warm_kill flu<br>heat disappear | I happen to think the warm weather is helping coronavirus not spread to the elderly in Arizona and Florida.<br>Some scientists think the virus will lose its effect as the weather gets warmer. Some oppose it. What do you think?<br>Why does there seem to be an acceptance of the theory that the virus will die off in warm weather? If that were the case, places like Thailand, Iran, and India would have fared much better. I think we'll see a 2nd wave in June because we opened too fast. | 3.97% |
| 13 (Temperature) | temperatur high humid degre<br>transmiss studi surviv no cold<br>affect research new find low<br>impact reduc human summer<br>know countri | Two interesting studies on coronavirus and weather. First shows worst-hit places have similar latitude, temperature and humidity. Based on travel routes, you'd expect SE Asia to be badly affected, but situation in northern green area far more severe<br>It's summer here, looks like #coronavirus is stronger on cold temperatures. Here in my city it's pretty warm weather.<br>Scientists and medical professionals have confirmed that the warm weather of the spring will be the end of the Coronavirus, and it cannot live in the amazingly warm and tropical temperatures that your cruise will be sailing to. | 3.65% |
| 14 (Responses to Trump February Comments) | disappear disappear_warm magic<br>magic_disappear april hoax<br>april_warm miracul democrat<br>miracl arriv presid come rememb<br>claim warm_april zero peopl right<br>tell | Have a look at Trump's tweet related to the COVID-19. 2 weeks ago he and his so called pandemic team claim very loudly that it will disappear with the warm weather. You've seen nothing yet and last time I checked God does not manufacture vaccines.<br>You're right, Trump was busy in February on the coronavirus response.<br>He was busy DOWNPLAYING it. Saying that 15 infections would soon be zero and that it would disappear in the warm weather. All in his words. | 4.75% |
| 15 (Warm Locations) | stop warm_stop stop_spread no summer outbreak expert hope help australia peopl new know singapor time count warm_help tell spring studi | How do you plan to deal with COVID19? Trump thinks the warmer weather will stop it, but with Singapore having 80F and the virus thriving despite the heat, I doubt he is right.<br>He clearly didn't factor in Singapore, whose warm weather hasn't stopped COVID-19 cases. He doesn't factor in much at all.<br>Summer may slow the growth of the coronavirus pandemic in the US but will NOT stop the spread... | 3.98% |
| 16 ('News') | news good bad help warm_kill humid know no research studi heat summer new warm_help peopl tell find die reduc stop | Bad news – warmer weather means higher UV index's which increase risks for skin cancer!<br>Good news, if true. South Asia Investor Review: Chinese Study: Hot and Humid Weather to Reduce Spread of #Covid_19<br>There goes the warm weather solves the COVID problem argument. If the warmer weather is having an impact, that is really, really bad news this fall and winter. | 1.64% |
| 17 ('Probably') | probabl away_warm stop slow summer season transmiss dr flu sar know no diseas right studi die read peopl think suggest | Since it appears than sunlight and warm humid weather kills COVID, these people were probably safer at the pool and golf course than they were in a cool, dry dark home anyway.<br>Will we be saved by summer in the northern hemisphere? Probably not.<br>He's so dumb he probably wants to hold out 'til April weather warms up, and then Covid-19 will magically go away. | 1.24% |
| 18 (Trump Criticism of Coronavirus Handling) | away_warm hoax tell think sar know peopl presid no flu time season democrat right april state rememb come claim look | When the first cases in China were reported we had 2-3 critical months to prepare for this and what did Trump do? He played golf, held rallies and downplayed COVID19 saying in his infinite wisdom that it will go away when the warm weather comes.<br>A prestigious scientific panel told the White House that it doesn't look like coronavirus will go away once the weather warms up.<br>@realDonaldTrump: A Hoax? Coronavirus will co away in warm weather? Do you ever listen to your hateful rhetoric? | 4.04% |
| 19 (Climate) | climat chang countri summer humid cold temperatur affect peopl condit slow help think pandem season differ current diseas singapor surviv | This might be fairly obvious by now, but COVID-19 has become established even in places with very warm climates.<br>Summer is approaching! Could warmer climates particularly in Africa stop the spread of #Coronavirus?<br>Pandemics like the coronavirus may occur more often when climate change is unabated. Warming and changing weather patterns shift the vectors and spread of disease. | 2.06% |
| 20 (Slow Spread) | slow warm_slow slow_spread hope scientist summer hope_warm studi help new warm_help pandem time stop research suggest transmiss novel spring countri | Interesting theory about warmer weather slowing down the spread of the Covid19.<br>The notion that warm weather will slow the coronavirus will be proven one way or the other in Houston in the next 2 weeks.<br>Warm weather won't slow the spread of COVID-19, but closing schools and maintaining physical distancing will, suggests a study led by St. Michael's Dr. Peter Juni and researchers. | 3.91% |
| 21 (Mocking Trump) | idiot april away_warm disappear think no believ peopl april_warm come democrat hoax presid know claim warm_april trump_warm countri heat away_april | Is anyone surprised that Donald Trump who is an idiot put Mike Pence who is an equal idiot in charge of the Coronavirus...<br>He took a gamble warmer weather would stop covid? Does the idiot know it has struck on both hemispheres of the globe?<br>Mike Pence and Donald Trump don't believe in global warming and yet they feel if the weather warms up it will kill the virus. Morons | 0.98% |
| 22 (Social Distancing) | distanc social social_distanc peopl summer effect no mask measur slow help research reduc stay need safe transmiss humid test high | With the warmer weather here, we remind all to maintain proper physical distancing. If you choose to go out, please respect physical distancing and say hello from a distance.<br>Let's hope we get a heat wave in April which looks like will reduce transmission of #covid-19 - we need it here in the Black Country as the increase in the number of cases is scary. Need hot weather and need social distancing to be taken seriously<br>Many think that when the weather warms up that Covid 19 will diminish. If so than why are warmer climates like Africa and | 1.09% |
|  |  | South America impacted. I think we will have to maintain social distancing until a vaccine is found. |  |
| 23 (Cold Weather) | snow cold_snow snow_kill cold new transmit myth fact area_hot transmit_area area diseases climat hot_humid no humid temperatur reason believ hand | There is no reason to believe that cold weather can kill the new coronavirus or other diseases.<br>WHO has published the fact that COVID-19 virus can be transmitted in areas with hot and humid climates. Neither Cold weather and snow CANNOT kill the new coronavirus.<br>Cold weather and snow CANNOT kill the new coronavirus. #coronavirus | 1.16% |
| 24 (Hot Weather) | surviv surviv_hot heat cold peopl no summer africa think degre know day tell temperatur believ long humid condit high countri | Everything in the summer should be gooch because COVID can't survive above 80 degree weather!<br>May be or may be not. Covid19 is here to stay. India with BCG vaccines, hot weather and immune system that withholds street food and drinking tap water, may survive better.<br>As hot as Nigeria is, coronavirus won't survive becassue it doesn't like hot weather. | 2.72% |
| 25 (Humidity) | humid warm_humid hot_humid slow studi heat transmiss summer research help high new slow_spread countri condit hope signific find temp singapor | #PublicHealth measures and not hot weather will help control the #pandemic<br>Hotter, humid weather may not halt spread of COVID-19<br>"Though some viral illnesses seem to slow in the summer months, this isn't always the case. In countries such as Australia and Iran, COVID-19 has spread very quickly despite warm and humid weather." | 4.15% |

\*Clusters generated using K-Means Algorithm

**Table S9:** Clustering Proportions by Algorithm; each algorithm clustered the relevant corpus into ten topics, and the proportion allocated to each topic is indicated below. Cluster numbers are not indicative of topic numbers, and are sorted in descending order.

|  | **k-Means** | **k-Medoids** | **LDA** |
| --- | --- | --- | --- |
| Median | 2.47% | 1% | 0% |
| 1 | 38.29% | 81% | 100% |
| 2 | 5.81% | 2% | 0% |
| 3 | 4.75% | 2% | 0% |
| 4 | 4.15% | 1% | 0% |
| 5 | 4.04% | 1% | 0% |
| 6 | 3.98% | 1% | 0% |
| 7 | 3.97% | 1% | 0% |
| 8 | 3.91% | 1% | 0% |
| 9 | 3.65% | 1% | 0% |
| 10 | 3.56% | 1% | 0% |
| 11 | 3.21% | 1% | 0% |
| 12 | 2.72% | 1% | 0% |
| 13 | 2.47% | 1% | 0% |
| 14 | 2.06% | 1% | 0% |
| 15 | 1.67% | 1% | 0% |
| 16 | 1.67% | 1% | 0% |
| 17 | 1.64% | 1% | 0% |
| 18 | 1.28% | 1% | 0% |

|  | ***k*-Means** | ***k*-Medoids** | **LDA** |
| --- | --- | --- | --- |
| 19 | 1.26% | 1% | 0% |
| 20 | 1.24% | 0% | 0% |
| 21 | 1.16% | 0% | 0% |
| 22 | 1.09% | 0% | 0% |
| 23 | 1.09% | 0% | 0% |
| 24 | 0.98% | 0% | 0% |
| 25 | 0.35% | 0% | 0% |

## Notes

### Competing Interest Statement

The authors have declared no competing interest.

### Author Declarations

Not applicable; no human was involved in this study.

